# A Deep Autoencoder for Fast Spectral-Temporal Fitting of Dynamic Deuterium Metabolic Imaging Data at 7T

**DOI:** 10.1101/2025.09.09.25335269

**Authors:** Aaron Paul Osburg, Amirmohammed Shamaei, Bernhard Strasser, Fabian Niess, Anna Duguid, Viola Bader, Sabina Frese, Lukas Hingerl, Hauke Fischer, William T. Clarke, Georg Langs, Wolfgang Bogner, Stanislav Motyka

## Abstract

Deuterium metabolic imaging (DMI) is a non-invasive magnetic resonance spectroscopic imaging technique enabling *in vivo* mapping of glucose metabolism. Dynamic DMI provides time-resolved metabolite maps and allows spatially resolved fitting of metabolic models to capture metabolite concentration dynamics. However, conventional fitting tools often require long processing times for high-resolution datasets, limiting their practical utility.

To address this bottleneck, we propose a deep autoencoder (DAE) for rapid spectral–temporal fitting of dynamic DMI data, supporting arbitrary parametric model constraints to describe metabolite concentration dynamics. The DAE was benchmarked against spectral-temporal fitting using FSL-MRS and LCModel. Fitting accuracy was evaluated on *in vivo* and synthetic whole-brain dynamic DMI data acquired at 7T using Bland-Altman metrics, Pearson correlation coefficients, structural similarity index measures, and root mean squared errors for both metabolite concentrations and model constraint parameters.

The DAE achieved processing times of 0.29 ms per voxel, corresponding to an acceleration of more than three orders of magnitude compared to LCModel/FSL-MRS (0.55/0.65 s per voxel). On *in vivo* data, it showed excellent agreement with LCModel, and on synthetic data, it consistently outperformed both reference methods across all evaluation metrics.

The proposed DAE enables accurate spectral-temporal fitting for whole-brain dynamic DMI scans within less than a second, matching or exceeding the performance of conventional state-of-the-art fitting methods. This makes it a promising tool for integration into efficient post-processing pipelines for research and clinical DMI workflows.

## 1. Introduction

Glucose (Glc) is the primary energy source for neuronal activity, neurotransmitter release, and brain cell communication in the mammalian brain (Dienel, 2019; Magistretti and Allaman, 2015). Understanding Glc metabolism of the human brain can provide insights into various neurodegenerative, epileptic, and psychiatric conditions, as well as cancer (Koppenol et al., 2011; Norat et al., 2020; Manji et al., 2012).

Deuterium metabolic imaging (DMI) is an emerging non-invasive magnetic resonance spectroscopic imaging (MRSI) technique enabling spatially and temporally resolved imaging of deuterium labeled substrates such as [6,6’]-^2^H-Glc and downstream metabolites like glutamate and glutamine (Glx) or lactate (Lac) (Lu et al., 2017; De Feyter et al., 2018; Ruhm et al., 2021; Serés Roig et al., 2023; Veltien et al., 2021). Due to the low natural abundance of deuterium (0.015 %), DMI spectra typically exhibit only a small number of well separated peaks and a relatively flat baseline, and the deuterated water (HDO) peak can serve as an internal concentration reference (Niess et al., 2024). However, DMI generally suffers from low signal-to-noise ratio (SNR) and long acquisition times, especially in dynamic ^2^H-MRSI experiments, which require multiple scans over time to capture metabolite concentration dynamics. While post-processing methods such as low-rank denoising (Nguyen et al., 2013) can partially mitigate this, resolution and SNR remain major limitations.

Spectral fitting is essential to determine the metabolite concentrations from MRSI spectra. There are multiple established fitting tools using various approaches to spectral fitting such as LCModel, ProFit1D, and FSL-MRS (Provencher, 1993, 2001; Borbath et al., 2021; Clarke et al., 2021, Clarke et al., 2024). Due to its long-standing, broad adoption and its robustness, LCModel is often regarded as the de-facto gold standard *in vivo* magnetic resonance spectroscopy (MRS) analysis software (Soher et al., 2022), especially for proton (^1^H) MRS. It performs spectral quantification by fitting a linear combination of basis spectra combined into a basisset, where each relevant metabolite is represented by one basis spectrum, most commonly generated using quantum mechanical simulations. However, spectral fitting with conventional tools is time-consuming, limiting the clinical applicability of (dynamic) DMI. For example, a whole-brain dynamic DMI scan may contain more than 10^4^ individual spectra, requiring over ten minutes for quantification with LCModel.

To reduce fitting time, massive GPU parallelization and deep learning (DL-) based approaches have been proposed for ^1^H-MRS(I) data (Gurbani et al., 2019; Shamaei et al., 2023, 2025; Giuffrida et al., 2025; Turco et al., 2024). These DL approaches, once trained, offer rapid inference and have demonstrated substantial speedups.

For dynamic DMI, fitting metabolic models that describe the concentration dynamics can provide additional valuable information compared to fitting each time point independently. The conventional spectral-temporal fitting approach performs separate, one dimensional spectral fits per time point, followed by temporal modeling of the resulting concentration curves, and is referred to as *1D fitting*. In contrast, FSL-MRS enables simultaneous spectral-temporal modeling of all spectra in a time course within a single fitting process, which was shown to result in improved fits, and is referred to as *2D fitting* (Tal, 2023; Clarke et al., 2024; Frese et al., 2024). However, long processing times remain a limitation of 2D fitting with FSL-MRS.

In this work, we propose a deep autoencoder (DAE) for fast 2D fitting of dynamic DMI data. The model combines a convolutional neural network (CNN) encoder with a physics-informed model decoder that learns to fit spectra as a linear combination of metabolite basis signals, building on earlier work by Shamaei et al. (2023, Shamaei et al. 2025). The decoder incorporates physical prior knowledge about the signal formation, thereby enforcing a physically interpretable latent space that includes metabolite concentrations and their dynamics. This approach is combined with a 2D fitting scheme enabling arbitrary parametric model constraints for the concentration dynamics. The proposed model was trained and evaluated on *in vivo* and synthetic DMI data and benchmarked against the 2D fitting tool of FSL-MRS and LCModel-based 1D fitting. To the best of our knowledge, this is the first DL-based approach applied to and developed specifically for quantification of DMI data and for 2D fitting of dynamic MRS data.

## 2. Materials and Methods

Details about the architecture of the proposed DAE are given in Section 2.1, while Sections 2.3 and 2.4 describe the *in vivo* and synthetic data used for training and evaluation. Sections 2.2 and 2.5 outline the evaluated model configurations and the training procedure. The methods used to assess the model performance are described in Section 2.7.

### 2.1. Network Architecture

A DAE is a neural network architecture commonly used for dimensionality reduction and feature learning (Goodfellow et al., 2016; Alain and Bengio, 2014). It consists of two subnetworks referred to as encoder ℰ and 𝒟 decoder, where a forward pass corresponds to the composition 𝒟°ℰ. The encoder maps the input to a latent representation, which the decoder transforms into its output, referred to as the reconstruction of the original input. The network is trained by minimizing a loss function *L* that quantifies the discrepancy between the input and its reconstruction (e.g., mean squared error), defining a self-supervised learning task. Ideally, this results in an encoder that has learned to extract useful features from the inputs. In conventional settings, DAEs aim to learn compact or descriptive latent features without a predefined interpretation. In contrast, our application focuses on learning latent representations that correspond to physically interpretable quantities, including metabolite concentrations and their temporal evolution.

Therefore, we propose a modified DAE architecture tailored to the spectral-temporal fitting task (see Figure 1). Its encoder is a CNN that processes sequences of complex-valued spectra dynamically acquired over time at a fixed location (spectral time courses), i.e., the inputs are two-dimensional samples with a spectral and a temporal axis, where spectra acquired at successive time points are stacked along the temporal axis. The decoder part is non-trainable and implements a forward physical signal model that describes spectral time courses as a function of multiple physical input parameters, e.g., accounting for metabolite concentration dynamics, frequency shift, and lineshape effects (see Sections 2.1.2 and 2.1.3 for a detailed description of the decoder and encoder).

**Figure 1:**
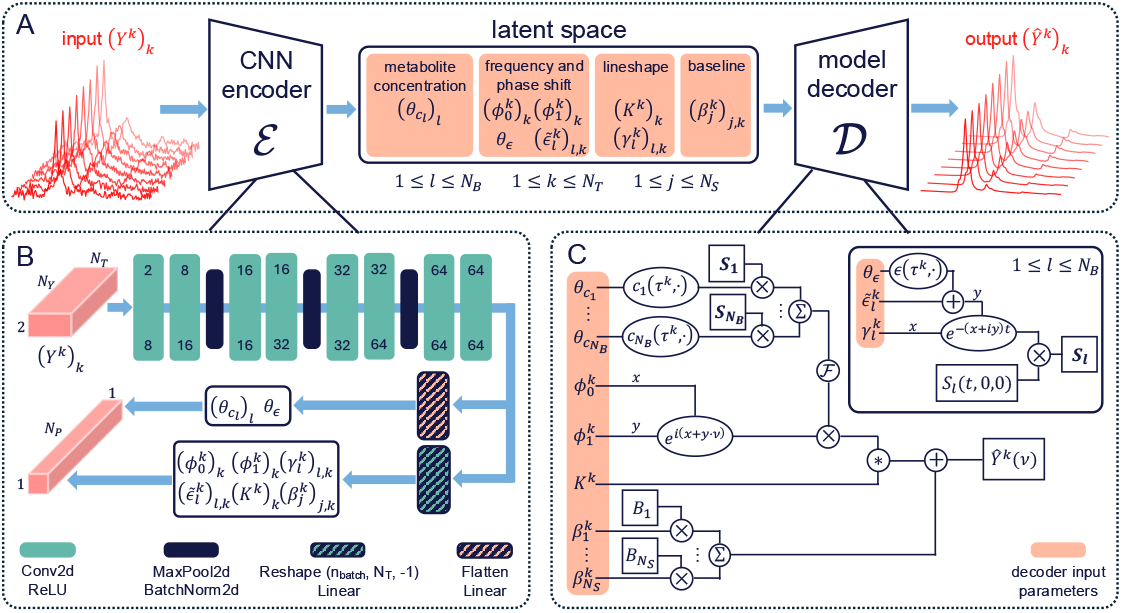
Overview of the proposed DAE architecture. (A): Forward pass of a spectral time course through the DAE. Latent space variables are interpretable as physical parameters in the signal model defined by Equation (1). (B): Architecture of the CNN ℰ encoder . The quantity *N*_*Y*_ denotes the number of points along the spectral axis and *N*_*P*_ the number of encoder output parameters per spectral time course. The frames representing convolutional layers show the number of input/output channels at the top/bottom. Latent variables for the lineshape kernel are passed through a softplus function and normalized according to Equation (5) before being passed to the decoder 𝒟 . (C): Computational graph implementing the signal model of 𝒟.

During training, the encoder learns to extract parameter values that allow the decoder to reconstruct the input signal as accurately as possible. We assumed that this is the case if and only if the features extracted by the encoder correspond to the physical input parameters of the decoder. After training, new samples to be quantified are passed through the encoder, yielding latent representations that contain parameters describing the metabolite concentration dynamics.

#### 2.1.1. Preprocessing

To ensure consistency of the DAE inputs, each spectral time course undergoes a preprocessing pipeline before being shown to the model. After zero-padding to a length of 256 in the time-domain, each spectrum of the time course is rotated in the complex plane such that the point of maximum absolute value lies on the real axis (i.e., its imaginary part vanishes). For DMI data, this point typically corresponds to the center of the HDO peak, making this step a naive, preliminary zero-order phase correction based on the HDO signal. Next, the signals are cropped to the frequency range between 0.7 ppm and 6.7 ppm. Finally, each spectral time course is normalized via z-scoring, i.e., the signal is mean-centered and divided by its standard deviation, computed jointly over the spectral and temporal dimensions. All preprocessing steps are cached, and used to rescale the quantification results, ensuring that the fits are consistent with the original signal amplitudes before normalization.

#### 2.1.2. Physics-Informed Decoder

Each spectrum in a time course with *N*_*T*_ spectra acquired at times 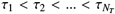 is represented by the decoder as the sum of a linear combination of *N*_*B*_ basis signals and a cubic B-spline baseline. The decoder model for the *k*-th spectrum 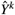 can be expressed analogously to LCModel’s signal model (Provencher, 1993) as

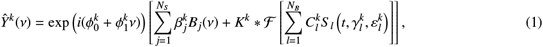

where *i* is the imaginary unit, 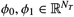 are the zero- and first-order phase corrections and ℱ represents the discrete Fourier transform. The *l*-th time-domain basis signal with concentration 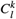 is denoted *S* _*l*_(*t*, 0, 0), and is broadened with the Lorentzian lineshape parameter 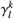 as well as frequency shifted by 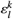,i.e.,

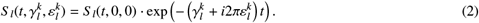

The time evolution of the concentration levels and frequency shift is modeled with parametric model constraints *c*_*l*_, *ϵ*, in other words

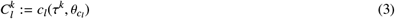

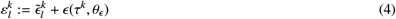

where *θ*_*cl*_, *θ*_*ϵ*_ are the model constraint parameters, and the model constraints may be any Python functions compatible with automatic differentiation in PyTorch. The quantities 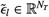 and 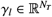 allow for slight differences between the frequency shifts and line broadenings of the basis signals at each time point. Penalties on these parameters are introduced in the DAE’s training loss to ensure these variations in fact remain small (see Section 2.2). The linear combinations of basis signals are convolved in the frequency domain with model-free lineshape kernels 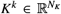 with *N*_*K*_ components, subject to the normalization constraint

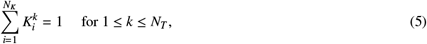

accounting for any peak distortions occurring *in vivo*, for example caused by *B*_0_-inhomogeneities (Provencher, 1993; Nguyen et al., 2013).

The baseline is modeled with a basis of *N*_*S*_ cubic B-splines, where the *j*-th basis function with weight 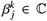 is denoted *B*_*j*_. Splines are used to represent the real and imaginary part of the baseline, thus the weights are complex-valued. The knots of the B-spline basis are distributed equidistantly over the spectral range, with one additional knot placed outside each boundary, and no boundary conditions are imposed. Smoothness of the baseline and lineshape kernel is imposed via regularization terms in the DAE’s loss function (see Section 2.2).

LCModel’s signal model is largely consistent with Equation (1), but models only one spectrum at a time instead of a full spectral time course simultaneously, which prevents imposing model constraints along the temporal axis. Furthermore, LCModel allows the components of the lineshape kernel to take negative values, while the *K*^*k*^ in Equation (1) are restricted to positive values.

The decoder used simulated ^2^H-resonances of HDO (4.8 ppm), Glc (3.9 ppm), Glx (2.4 ppm) and Lac/fat (1.3 ppm) as a basisset (Naressi et al., 2001; Starčuk et al., 2009). Internally, all decoder inputs are scaled by heuristic constants (see Supporting Information Table S1) to keep typical values around unity, which improves training speed.

#### 2.1.3. Encoder

The encoder of the proposed DAE follows a classical CNN architecture with two input channels representing the real and imaginary parts of the input spectral time courses. It consists of four convolutional blocks (see Figure 1B), each comprising two 2D convolutional layers with the ReLU activation function. Max-pooling and batch normalization are applied between consecutive blocks. The final convolutional block is followed by two parallel linear layers. The first is used to infer parameters, which are fitted independently at each time point (e.g., phase correction and baseline parameters). For this purpose, the features are reshaped to a three dimensional tensor with size *N*_*T*_ along the second dimension. The second one, preceded by a flattening operation, estimates the remaining parameters, which require context over the full spectral time course, such as model constraint parameters describing the concentration dynamics. All convolutional layers used a kernel size of (3,5) and padding of (1,2). In the last block, the stride was set to (1,2), in all remaining blocks, it was (1,1). A complete overview of the architectural parameters is provided in Supporting Information Table S2. Parameters subject to a (physical) non-negativity constraint such as metabolite concentrations, or the components of the lineshape kernels, are additionally passed through a softplus function. The encoder ensures the fulfillment of the normalization constraint of the lineshape kernel by dividing the corresponding output components by their sum.

### 2.2. Loss Function

The loss L minimized during DAE training consists of a squared *L*^2^-norm penalty on the complex reconstruction residuals and multiple regularization terms

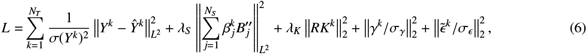

where *Y*^*k*^ denotes the *k*-th spectrum of a training sample (i.e., one spectral time course), Ŷ^*k*^ is its reconstruction, and 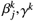, and 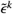 are the encoder predictions for the corresponding parameters from Equations (1) and (4). Here, 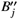 denotes the second derivative of the *j*-th spline basis function (approximated with finite differences), *R* is a second-derivative regularization matrix as defined in Equation (3.12) in Provencher (1982), and ∥.∥_*L*2_, ∥.∥_2_ denote the (discrete) *L*^2^- and 2-norms, respectively. The second and third terms in Equation (6) impose smoothness on the baseline splines and the lineshape kernels, and the remaining regularization terms enforce component-wise small values of *γ*^*k*^ and 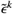 for each *k*.

The contribution of each term is weighted using the *σ*(*Y*^*k*^), denoting the standard deviation of the noise in *Y*^*k*^, and the hyperparameters *λ*_*S*_, *λ*_*K*_, *σ*_*γ*_, *σ*_*ϵ*_ . Here, *σ*(*Y*^*k*^) was approximated by the empirical standard deviation of the reconstruction residuals. During training, *σ*(*Y*^*k*^) was treated as a constant in each update step, i.e., its dependence on *Ŷ*^*k*^ (and thus on the DAE’s trainable parameters), was not considered when computing gradients.

It is important to note that all terms in Equation (6) depend solely on the input data, their reconstruction, and the encoder output. Consequently, the DAE can be trained in a self-supervised manner using only a set of spectral time courses − and does not require ground truth (GT) data, which is particularly advantageous when working with *in vivo* data, where GT concentrations are inaccessible.

The loss function strongly resembles the cost function of the nonlinear least-squares spectral fitting problem solved by LCModel. However, in LCModel the weights of the regularization terms *λ*_*S*_ and *λ*_*K*_ may be varied during spectral fitting, while they were kept fixed during the training of the proposed DAE. LCModel only considers the real part of the spectrum in the least-squares fit, which was shown to be unfavorable for the quality of the fits (Sokolenko et al., 2019). Therefore, both the real and the imaginary parts are considered for the calculation of the reconstruction loss and baseline spline regularization in Equation (6).

### 2.3. In Vivo Dynamic DMI Data

*In vivo* dynamic DMI data were acquired from six healthy volunteers (age: 26 ± 2 years; BMI: 23 ± 4 kg/m2, 4 male/2 female) in a previous study by Niess et al. (2024). All participants underwent overnight fasting and oral administration of [6,6’]-^2^H-Glc (0.8 g/kg body weight, ≥ 99 % purity, Cambridge Isotopes, USA) dissolved in 200 ml water prior to scanning. The labeled glucose was consumed on the patient table of the MR scanner within one minute, right before the participants were moved inside the magnet bore.

All scans were performed on a Magnetom dot Plus 7T whole-body MR scanner (Siemens Healthineers, Germany) using a ^2^H/^1^H dual-tuned quadrature birdcage head coil (Stark Contrasts MRI Coils Research, Germany). Each subject underwent eight whole-brain DMI scans (*N*_*T*_ = 8), starting 14 minutes after glucose intake, with a temporal resolution of 7 minutes between scans (i.e., the individual scans were obtained at *τ*_*i*_ = (*i* + 1) · 7 min, 1 ≤*i* ≤*N*_*T*_, see Figure 2A). The study was approved by the local ethics committee of the Medical University of Vienna (EK No. 1767/2022, date of approval: 01/13/2025) and followed the guidelines of the Declaration of Helsinki.

**Figure 2:**
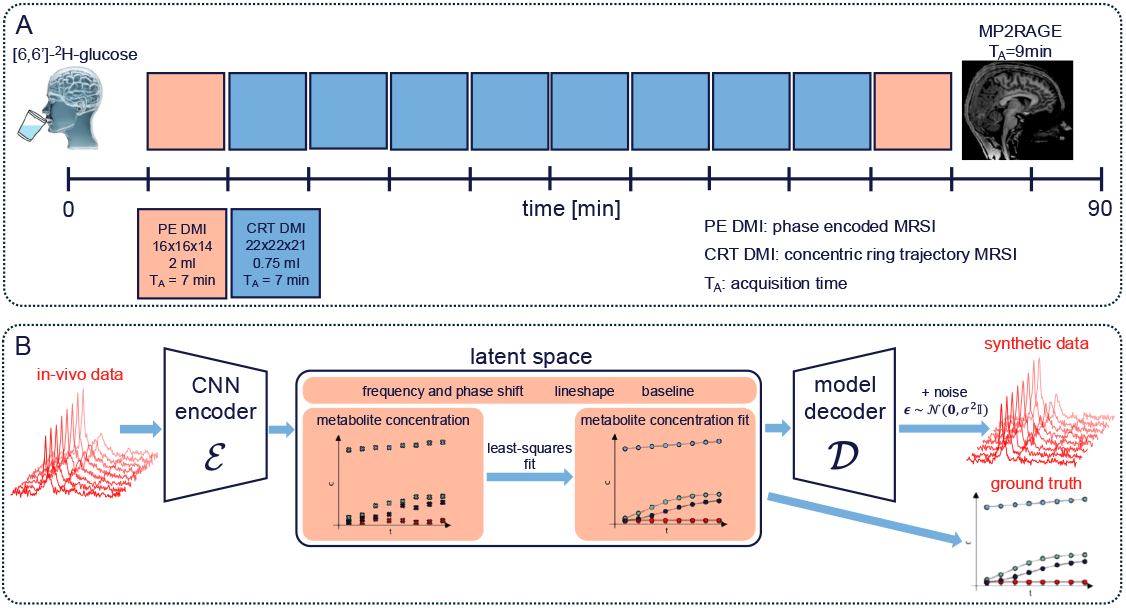
Data acquisition procedures for the *in vivo* (A) and the synthetic (B) data. (A): Schematic of the dynamic DMI experiment from Niess et al. (2024): After oral administration of [6,6’]-^2^H glucose and initial preparatory scans, ten DMI brain scans were acquired over ∼ 70 minutes using elliptical phase encoding (coral) or CRT readout (blue), followed by a T1-weighted image. The datasets used in this work consist of the central eight CRT-DMI scans. (B): Schematic of the generation of synthetic data from *in vivo* data: A DAE encoder trained on *in vivo* data 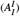 estimated independent per-time-point metabolite concentrations. Model constraints were then fitted and used to overwrite the original concentration estimates in the latent space. The modified vectors were re-injected into the decoder to synthesize spectral time courses with known dynamics, followed by Gaussian noise addition and inversion of cached preprocessing operations.

The data was acquired using a ^2^H free induction decay (FID) MRSI sequence with 3D density-weighted concentric ring trajectory (CRT-DMI) readout with 0.75 ml isotropic resolution and a non-localized rectangular excitation pulse (flip angle: 86^°^, pulse duration 500 *µ*s), TR = 290 ms, acquisition delay: 2.0 ms, matrix: 22 x 22 x 21, FOV: 200 x 200 x 192 mm3, *N*_*rings*_ = 43, bandwidth 380 Hz, number of samples 96) (Hingerl et al., 2018; Steel et al., 2018; Clarke et al., 2023; Chiew et al., 2018).

The CRT-DMI data were reconstructed using non-Cartesian 3D discrete Fourier transformation and global spatiotemporal low-rank denoising (rank 8) was applied to the reconstructed time-domain signals (see Niess et al. (2024) for details). Each subject’s scans were concatenated into a 5D array (three spatial, one temporal and one spectral dimension). Spectral time courses outside the brain were replaced with zeros.

Data of four subjects were used to create datasets for training (6666 spectral time courses) and validation (741 spectral time courses), and two subjects were retained for testing (1771 and 1902 spectral time courses). The average SNR across all spectra was 16 ± 5. In each voxel, SNR was defined as the ratio of the amplitude of the HDO peak to the standard deviation of noise in the spectral region above 6.7 ppm.

### 2.4. Synthetic Dynamic DMI Data

The *in vivo* data served as the basis for generating synthetic dynamic DMI data, which have known metabolite concentration dynamics that obey parametric model constraints, and which closely resemble *in vivo* data. This allowed direct comparisons of model constraint parameter and metabolite concentration fits with GT based on data with realistic spatial, spectral, and temporal features.

Initially, one DAE instance (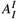, specified in Section 2.5) was trained with the *in vivo* data (see Supporting Information Figure S1 for representative spectral fits and metabolite maps compared to LCModel fits of the same data). This instance independently fits each spectrum of an input spectral time course (i.e., no model constraints were imposed on metabolite concentration dynamics). The trained DAE was used to create a synthetic “twin” for the *in vivo* dynamic DMI data of each subject, as illustrated in Figure 2B.

For each subject, the *in vivo* data was passed through the encoder of 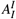, yielding latent representations including metabolite concentration estimates. Each concentration time series was subsequently fitted to parametric models using least-squares fitting with SciPy (Virtanen et al., 2020). A detailed description of the implementation is provided in Supporting Information Text S1 and Supporting Information Figure S2. The parametric models were chosen heuristically based on observed metabolite concentration dynamics, using logistic functions for Glc and Glx (Verhulst, 1838),

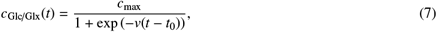

linear functions for HDO

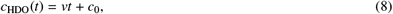

and constant functions for Lac

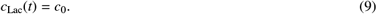

The fitted model constraints were then used to overwrite the encoder’s original concentration estimates in the latent representations before reconstructing the spectra. Gaussian noise was added to the reconstructed spectral time courses to achieve an average SNR slightly below that of the *in vivo* data. The cached preprocessing steps described in Section 2.1.1 were subsequently inverted, and the signals were truncated in the time-domain to the original FID length of 96 points before zero-padding. Finally, the synthetic data was reshaped into the original 5D array structure.

The resulting synthetic data was split into training, validation and test datasets in the same way as the in-vivo data. Importantly, the synthetic datasets contain GT concentration levels and model constraint parameters for each spectral time course. The average SNR across all synthetic data was 11 ± 3.

### 2.5. Implementation Details and DAE Training

All training runs and performance measurements were conducted on a computer with an AMD Ryzen 9 5950X 16-core CPU with 64 GB CPU RAM and one Nvidia GeForce RTX 4080 SUPER GPU with 16 GB GPU RAM, running Ubuntu 24.04.1 LTS. Models were implemented in Python 3.11.8 using PyTorch 2.3.1 and Pytorch Lightning 2.3.1 with CUDA 12.6. Training was performed for 800 epochs using the Adam optimizer (Kingma and Ba, 2014) with a batch size of 256, an initial learning rate of 0.002, and a weight decay of 0.001. The learning rate was halved at epochs 20, 50, 100, 200, 300, 400 and 600.

The lineshape kernel size was set to *N*_*K*_ = 16 (approximately 0.49 ppm), and the weight of the corresponding regularization term to *λ*_*K*_ = 30. Due to the characteristically flat baseline in DMI data, the number of spline knots was set to *N*_*S*_ = 4 with a strong baseline regularization of *λ*_*S*_ = 10^10^. The remaining regularization parameters *σ*_*γL*_ and *σ*_*ϵ*_ were set to values corresponding to 0.1 *s*^−1^ and 0.004 ppm, respectively. Hyperparameter values were determined based on LCModel default settings, heuristically based on properties of the DMI data (such as flat baselines and approximate linewidths), or found empirically.

Three DAE instances were trained for this study:

- 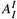: Trained on *in vivo* data and used to generate synthetic dynamic DMI data. No constraints on metabolite concentration dynamics were imposed (i.e., each time point was fitted independently by assigning *N*_*T*_ free parameters per metabolite).
- 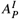: Trained on *in vivo* data with model constraints on concentration dynamics, as defined in Equations (7) to (9).
- 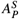: Trained on synthetic data with model constraints on concentration dynamics, as defined in Equations (7) to (9).

All other model and training parameters were identical across instances. Detailed settings are provided in Supporting Information Tables S1 and S2. No model constraints were applied to the frequency shift in any DAE instance.

### 2.6. Reference Standards

As *in vivo* metabolite concentrations and dynamics are unknown, 1D fitting using LCModel 6.3 was employed as the primary reference method due to its longstanding use in MRS analysis for more than three decades. Spectral fitting in LCModel was performed voxel-wise for each time point using an in-house post-processing pipeline for dynamic DMI data. Subsequently, model constraints were fitted to the resulting concentration estimates, analogously to the approach in Section 2.4. In the remainder of this work, “LCModel” refers to this 1D fitting method (including the model constraint fits), unless stated otherwise.

For additional benchmarking, the proposed method was also compared to the 2D fitting tool of FSL-MRS 2.3.1 (Clarke et al., 2024). This software was chosen because it was the only publicly available tool for 2D fitting of dynamic MRS data. Details of the LCModel and FSL-MRS configurations are given in Supporting Information Text S2.

### 2.7. Performance Analysis

#### 2.7.1. Validation with Synthetic Data

We evaluated the DAE instance 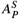 with respect to fitting accuracy using the synthetic test dataset. The performance of the proposed model and the reference methods was assessed via Pearson correlation coefficients (PCCs) of the fitted versus GT concentration levels for all time points as well as the root mean squared error (RMSE) and the structural similarity index measure (SSIM) (as defined in Supporting Information Text S3) between the corresponding metabolite maps and model constraint parameter maps. Because of different internal scalings, the parameters fitted by LCModel and FSL-MRS were rescaled to align with the GT data. For each of the methods, a global scaling factor was computed to minimize the RMSE between the fitted and GT HDO concentrations across both test subjects and all time points (see Supporting Information Text S4).

#### 2.7.2. Validation With in Vivo Data

For the *in vivo* test dataset, the fitting performance of DAE instance 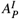 was analyzed using PCC, SSIM, and Bland–Altman plots to assess agreement with the primary reference LCModel. For comparison, the agreement between FSL-MRS and LCModel was assessed with the same metrics. LCModel and FSL-MRS fits were rescaled using the scaling factors obtained from the evaluations with synthetic data.

Mean processing times per spectral time course were measured for each method and both test subjects, using full CPU parallelization for FSL-MRS and LCModel, and GPU acceleration for inference with the DAE instances.

## 3. Results

### 3.1. Processing Times

Mean processing times per spectral time course/whole-brain dataset (𝒪(2000) spectral time courses) were approximately 0.3 ms/0.5 s for DAE models 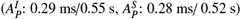, 0.55 s/17 min for LCModel, and 0.65 s/20 min for FSL-MRS (see Supporting Information Table S3 for per-subject values).

### 3.2. Performance on in Vivo Data

Metabolite maps from 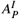 closely matched those of LCModel for the three major metabolite signals Glc, Glx, and HDO with respect to spatial features and signal intensity (see Figure 3A and Supporting Information Figure S3A). Noticeable differences exist for Glx at early time points, where LCModel maps exhibit relatively strong noise. For late time points, the Glc maps from 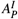 have a slightly elevated intensity compared to LCModel, while maintaining highly similar spatial features.

**Figure 3:**
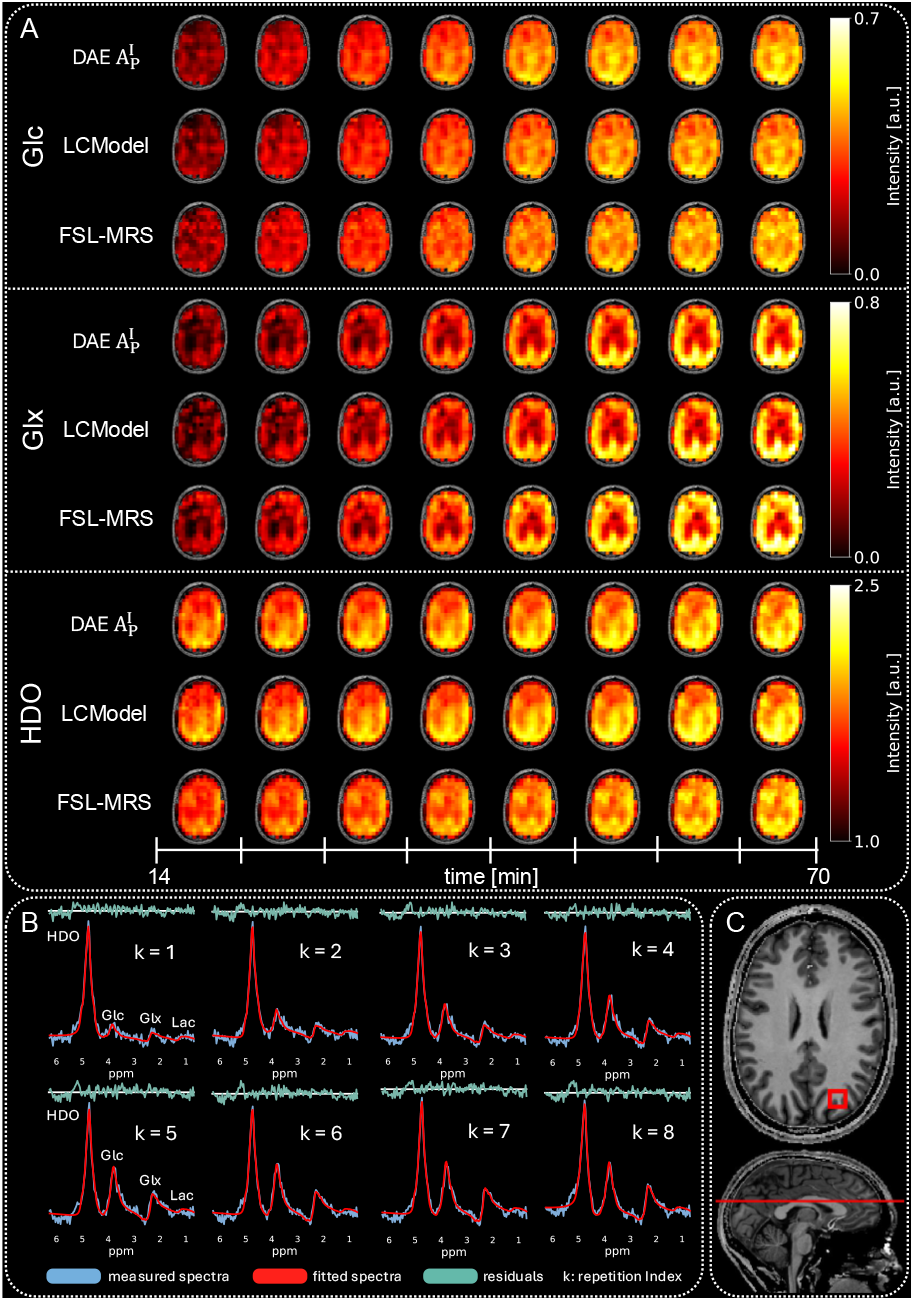
(A): Metabolite maps of the three major metabolite signals (Glc, Glx, HDO) across all eight time points for one subject from the *in vivo* test dataset, fitted by DAE instance 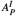 and the reference methods LCModel and FSL-MRS. (B): Example spectral time course with corresponding DAE fits and residuals. The index *k* indicates the position of each spectrum in the time series, and metabolite labels identify the spectral peaks. (C): T1-weighted anatomical image with a red box indicating the voxel location of the time course and a red line marking the position of the presented metabolite maps.

The visual similarity between FSL-MRS and LCModel/ 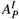 was lower for Glc and Glx, with noisier FSL-MRS maps (Glc in Figure 3A), clear deviations of the average intensity (Glx in Figure 3A), and slightly different spatial features (Glc maps for late time points in Figure 3A and Supporting Information Figure S3A). Strong differences were found between the HDO maps from LCModel/ 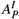 and FSL-MRS, showing lower overall concentration levels in the center region, resulting in altered spatial features. Lac maps, typically noise-dominated across all methods (see Supporting Information Figure S4), showed higher average signal intensities fitted by LCModel compared to 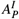 and FSL-MRS (LCModel: 0.062 a.u./0.051 a.u., FSL-MRS: 0.048 a.u./0.029 a.u.,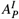: 0.040 a.u./ 0.030 a.u. for test subject 1/2).

Figure 3B shows the measured spectra, spectral fits from 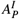, and their residuals for one representative spectral time course. Spectral fits from 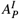 aligned well with measured spectra, with residuals distributed approximately symmetrically around zero. No dependency of the fit quality on the position of the spectrum in the time course was observed.

In Figure 4, the Glc, Glx, and HDO maps for one subject from the *in vivo* test dataset fitted by 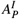 are shown alongside the corresponding model constraint parameter maps (analogous maps for the second subject are presented in Supporting Information Figure S5). The time shift parameter (*t*_0_) maps for Glc and Glx were spatially homogeneous, with consistently smaller values for Glc (average *t*_0_ for test subject 1/2: 22 ± 4 min/25 ± 3 min) than for Glx (average *t*_0_ for test subject 1/2: 30 ± 4 min/32 ± 3 min), suggesting a delayed signal onset for the downstream metabolite Glx. Visually, the *c*_*max*_ maps for Glc and Glx closely resembled the corresponding metabolite maps for the final time point. Analogously, HDO intercept maps (*c*_0_) showed high similarity to the early HDO metabolite maps. The slope parameter for HDO (*v*) was predominantly positive across the brain (mean *v* for test subject 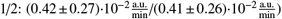, indicating a gradual increase of the HDO signal over time. These parameter maps provide interpretable summaries of dynamic signal changes and align well with physiological expectations.

**Figure 4:**
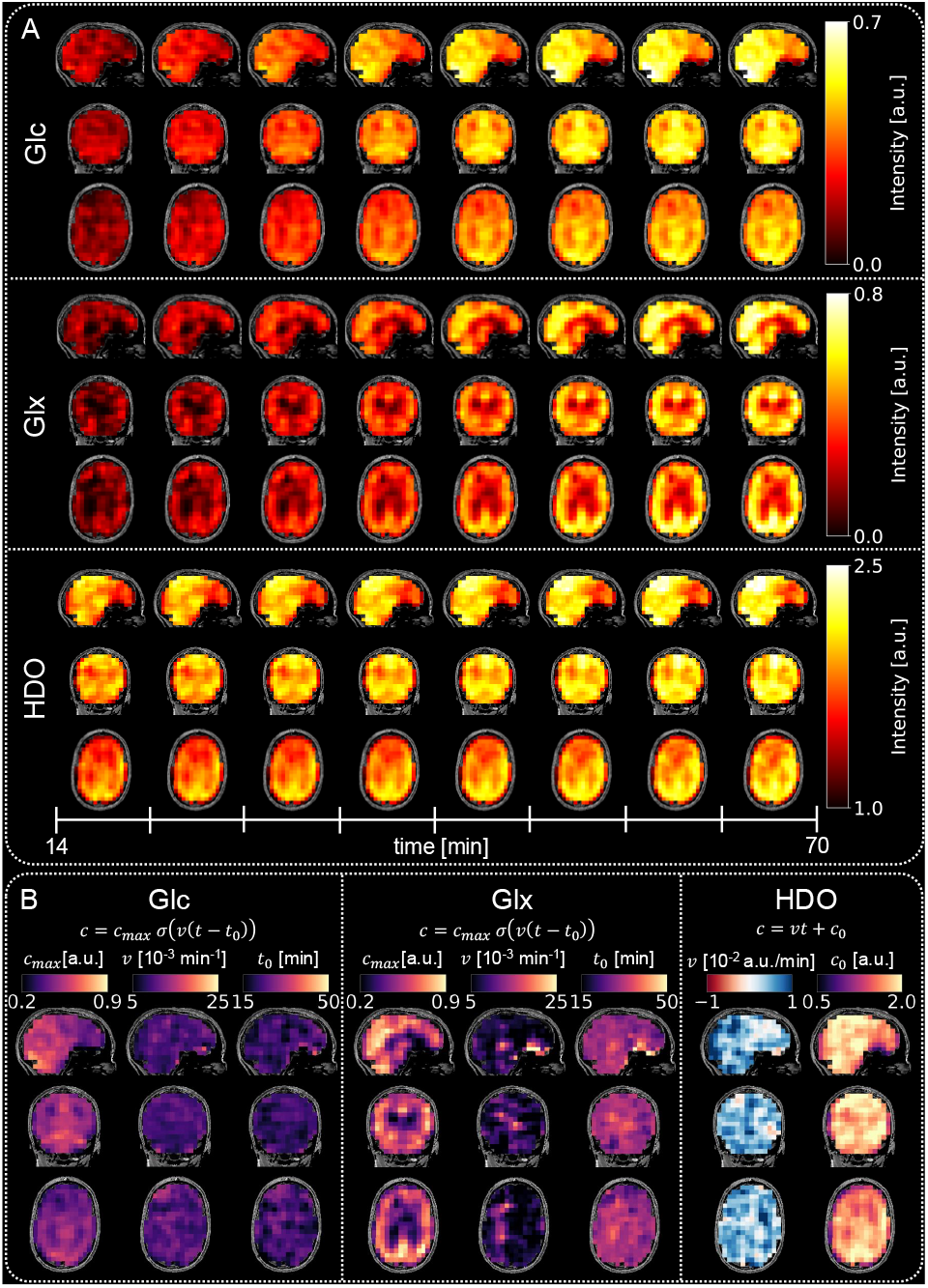
(A): Metabolite maps of the three major metabolite signals (Glc, Glx, HDO) for one subject of the *in vivo* test dataset and all eight time points fitted by DAE instance 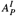 . For each metabolite, a sagittal (first row), coronal (second row) and axial (third row) map is shown. (B): Corresponding maps of the fitted model constraint parameters.

Bland-Altman plots for the concentration levels fitted by DAE instance 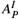 and FSL-MRS versus LCModel are presented in Figure 5. Corresponding Bland-Altman metrics, along with PCCs and SSIMs between 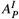 /FSL-MRS and LCModel are summarized in Table 1. Reported values are averaged across both test subjects (see Supporting Information Table S4 for per-subject metrics). The proposed DAE achieved very high structural similarity (SSIM ≥ 0.98) and strong correlation (PCC ≥ 0.94) for all high-SNR signals (Glc, Glx, HDO). In contrast, FSL-MRS yielded lower performance, with SSIMs ranging from 0.59 to 0.85 and PCCs between 0.39 and 0.81. For the low-SNR Lac signal, 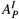 achieved a moderate SSIM and PCC of approximately 0.70. The percentage bias of 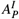 remained below 10% for Glc and Glx, and under 1% for HDO, and was consistently smaller than that of FSL-MRS. The Lac concentrations fitted by 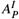 and FSL-MRS were on average lower than those fitted by LCModel, with biases of −48% and −46%, respectively. Across all metabolites, 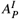 exhibited substantially narrower limits of agreement (LoA) than FSL-MRS (see Table 1 and Figure 5).

**Table 1:**
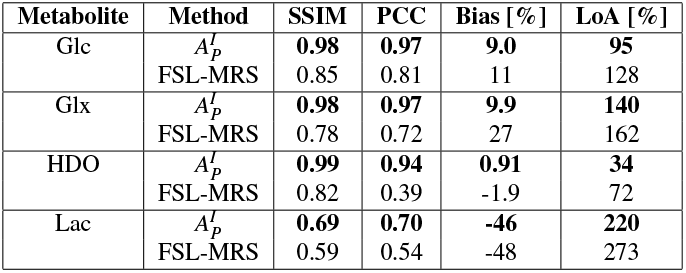
SSIM, PCC and Bland-Altman metrics (bias, LoA) for the metabolite concentration fits produced by DAE instance 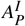 and FSL-MRS versus LCModel. The presented values are averages calculated over both subjects from the *in vivo* test dataset. Bias and LoA are expressed as percentages relative to the mean of the paired measurements.

| Metabolite | Method | SSIM | PCC | Bias [%] | LoA [%] |
| --- | --- | --- | --- | --- | --- |
| Glc | $A_p^I$ | **0.98** | **0.97** | **9.0** | **95** |
|  | FSL-MRS | 0.85 | 0.81 | 11 | 128 |
| Glx | $A_p^I$ | **0.98** | **0.97** | **9.9** | **140** |
|  | FSL-MRS | 0.78 | 0.72 | 27 | 162 |
| HDO | $A_p^I$ | **0.99** | **0.94** | **0.91** | **34** |
|  | FSL-MRS | 0.82 | 0.39 | -1.9 | 72 |
| Lac | $A_p^I$ | **0.69** | **0.70** | **-46** | **220** |
|  | FSL-MRS | 0.59 | 0.54 | -48 | 273 |

**Figure 5:**
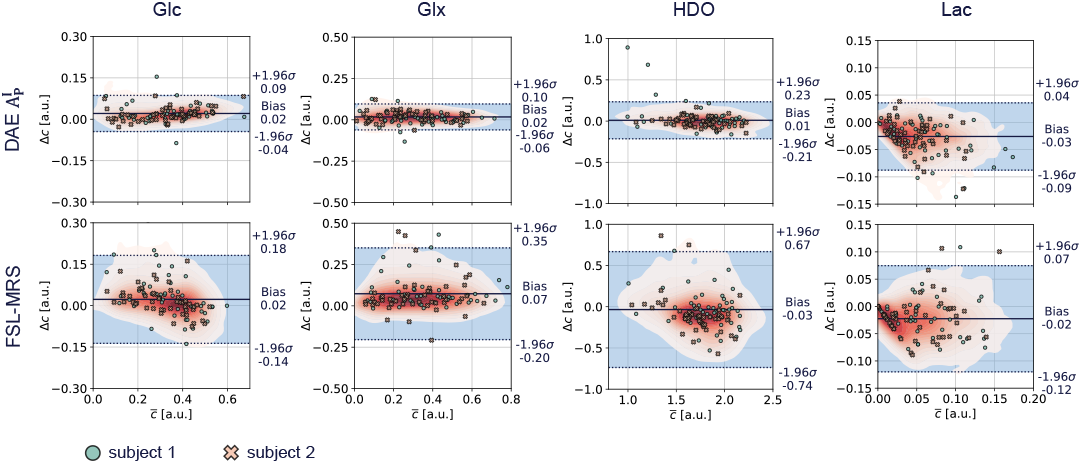
Bland-Altman plots of the metabolite concentrations fitted by DAE instance 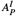 and FSL-MRS versus LCModel for the *in vivo* test dataset and all metabolite basis signals. The average of two compared methods is denoted by 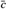, and their difference by Δ*c*. Data from both test subjects and all time points were included. Point distributions are visualized as density maps, with darker red indicating higher density. Additionally, 25 randomly sampled data points per subject are overlaid as markers.

### 3.3. Performance on Synthetic Data

Figure 6 shows the fitted metabolite maps from DAE instance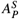, FSL-MRS, and LCModel as well as the GT maps for one subject from the synthetic test dataset at time points *k* = 2, 5, 8 (see Supporting Information Figure S6 for the second subject). The corresponding relative differences to the GT are shown alongside the fitted metabolite maps. The PCC, RMSE and SSIM values for the fitted metabolite concentration levels and model constraint parameters averaged over both test subjects are listed in Table 2 and Table 3 (see Supporting Information Tables S5 and S6 for per-subject metrics), correlation plots for the Glc model constraint parameters are shown in Figure 7 (analogous plots for Glx and HDO are presented in Supporting Information Figures S7 and S8). Lac was excluded from the model constraint evaluation since its dynamics were modeled with a constant function, making its model constraint parameters equivalent to the concentration fits.

**Table 2:** SSIM, PCC, and RMSE values for the metabolite concentration fits from 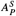, LCModel and FSL-MRS compared to the GT. The presented values are averaged over both subjects from the synthetic test dataset.

| Metabolite | Method | SSIM | PCC | RMSE [$10^{-2}$ a.u.] |
| --- | --- | --- | --- | --- |
| Glc | $A_p^S$ | **0.99** | **0.99** | **1.8** |
|  | LCModel | 0.97 | 0.98 | 3.2 |
|  | FSL-MRS | 0.96 | 0.94 | 4.6 |
| Glx | $A_p^S$ | **0.99** | **0.99** | **2.7** |
|  | LCModel | 0.98 | 0.98 | 4.0 |
|  | FSL-MRS | 0.97 | 0.97 | 4.1 |
| HDO | $A_p^S$ | **1.00** | **0.99** | **4.3** |
|  | LCModel | 0.97 | 0.95 | 9.9 |
|  | FSL-MRS | 0.96 | 0.84 | 16 |
| Lac | $A_p^S$ | **0.87** | **0.88** | **1.9** |
|  | LCModel | 0.83 | 0.85 | 2.3 |
|  | FSL-MRS | 0.79 | 0.74 | 2.7 |

**Table 3:** SSIM, PCC, and RMSE values for the model constraint parameters fitted by 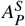, LCModel and FSL-MRS compared to the GT. The presented values are averaged over both subjects from the synthetic test dataset. Extreme outliers occurring in FSL-MRS and 1D fitting were removed using the modified z-score method with a threshold value of 5 before calculating the metrics (Iglewicz and Hoaglin, 1993).

| Metabolite | Parameter | Method | SSIM | PCC | RMSE [a.u.] |
| --- | --- | --- | --- | --- | --- |
| Glc | $c_{max}$ [a.u.] | $A_p^S$ | **0.99** | **0.98** | **0.020** |
|  |  | LCModel | 0.93 | 0.91 | 0.040 |
|  |  | FSL-MRS | 0.81 | 0.48 | 0.10 |
| | $v$ [$10^{-3}$ min$^{-1}$] | $A_p^S$ | **0.95** | **0.76** | **1.0** |
|  |  | LCModel | 0.82 | 0.50 | 2.4 |
|  |  | FSL-MRS | 0.62 | 0.40 | 3.4 |
| | $t_0$ [min] | $A_p^S$ | **0.98** | **0.91** | **1.7** |
|  |  | LCModel | 0.92 | 0.77 | 3.0 |
|  |  | FSL-MRS | 0.74 | 0.21 | 6.9 |
| Glx | $c_{max}$ [a.u.] | $A_p^S$ | **0.92** | **0.92** | **0.074** |
|  |  | LCModel | 0.77 | 0.81 | 0.11 |
|  |  | FSL-MRS | 0.85 | 0.81 | 0.11 |
| | $v$ [$10^{-3}$ min$^{-1}$] | $A_p^S$ | **0.87** | **0.74** | **2.0** |
|  |  | LCModel | 0.70 | 0.63 | 3.2 |
|  |  | FSL-MRS | 0.75 | 0.58 | 3.9 |
| | $t_0$ [min] | $A_p^S$ | **0.84** | **0.58** | **4.5** |
|  |  | LCModel | 0.78 | 0.46 | 7.5 |
|  |  | FSL-MRS | 0.80 | 0.40 | 6.7 |
| HDO | $c_0$ [a.u.] | $A_p^S$ | **0.90** | **0.88** | **0.14** |
|  |  | LCModel | 0.83 | 0.79 | 0.20 |
|  |  | FSL-MRS | 0.77 | 0.73 | 0.24 |
| | $v$ [$10^{-2}$ a.u./min] | $A_p^S$ | **0.99** | **0.97** | **0.068** |
|  |  | LCModel | 0.98 | 0.95 | 0.099 |
|  |  | FSL-MRS | 0.96 | 0.85 | 0.26 |

**Figure 6:**
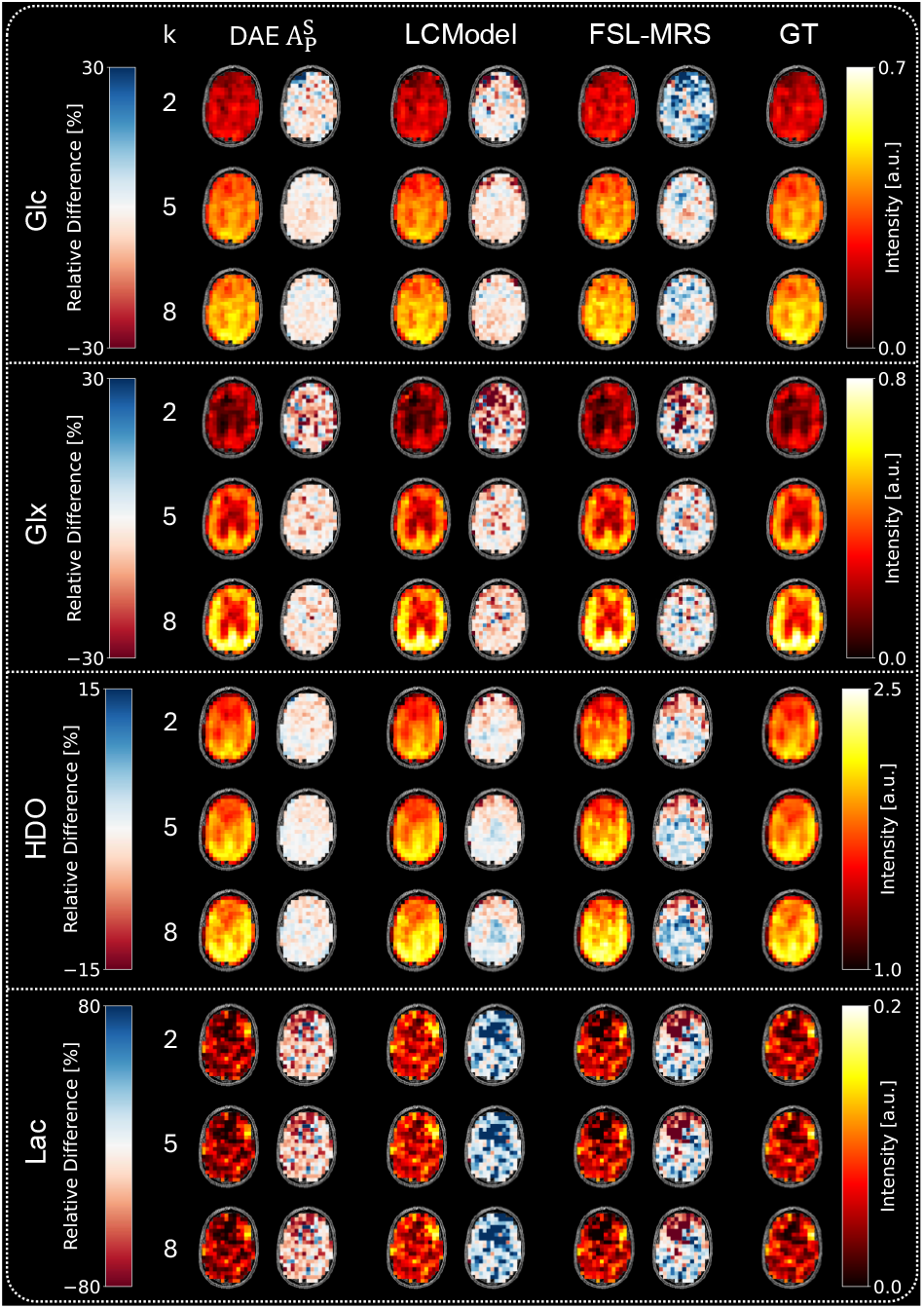
Metabolite maps of one subject of the synthetic test dataset for the time points *k* ∈2, 5, 8 from DAE instance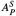, LCModel, and FSL-MRS. GT maps are shown in the right column. To the right of each metabolite map, a map of relative differences Δ*c*_*rel*_ = (*c* _*fit*_ *c*_*GT*_)*/c*_*GT*_ (where *c* _*fit*_ is the fitted and *c*_*GT*_ the GT concentration) to the GT is shown.

**Figure 7:**
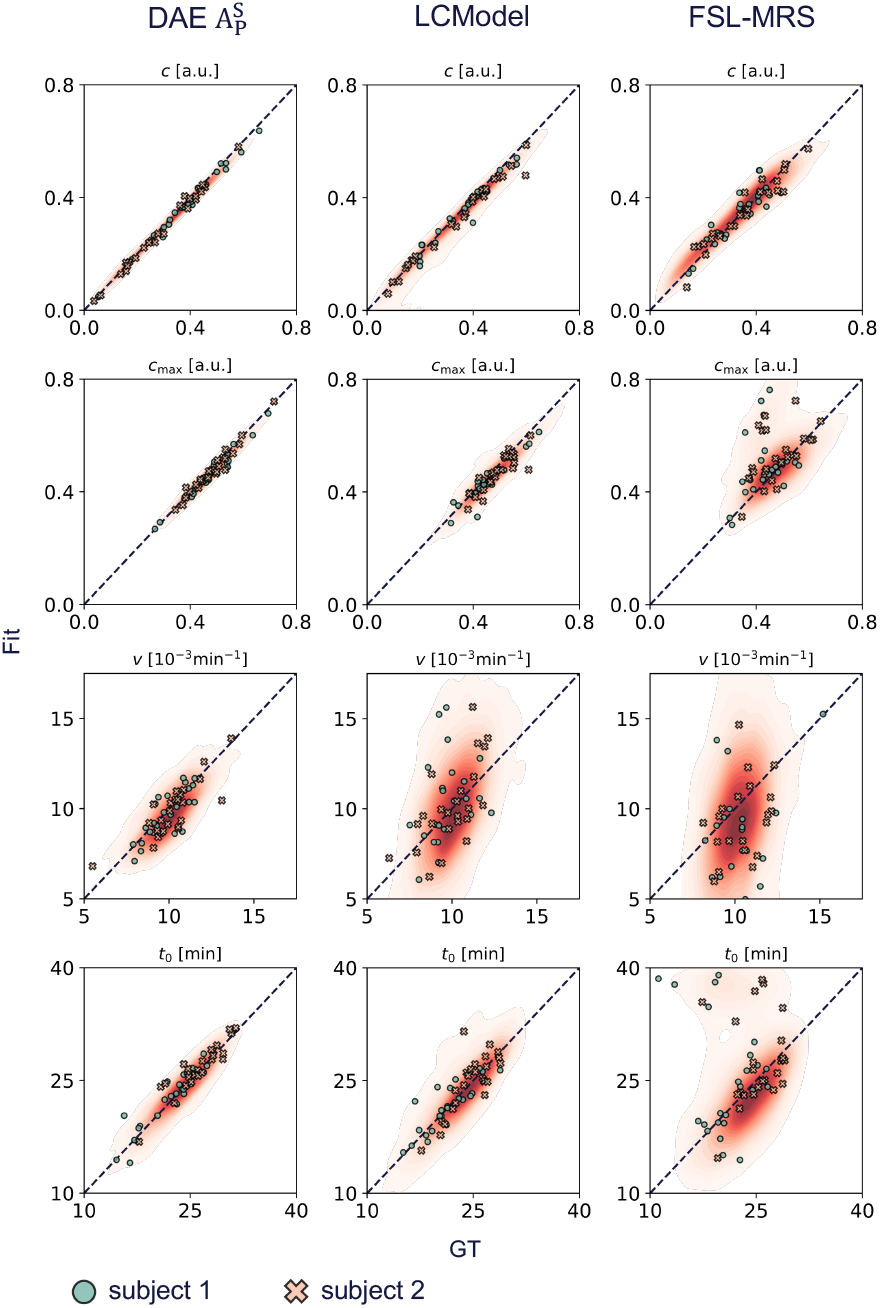
Correlation plot for the metabolite concentrations and model constraints for Glc from DAE instance 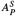, LCModel, and FSL-MRS versus GT for both subjects of the synthetic test dataset. Point distributions are visualized as density maps, with darker red indicating higher density. 25 randomly sampled data points per subject are overlaid as markers.

#### 3.3.1. Metabolite Concentration Maps

Visually, Glc, Glx, and HDO maps fitted with LCModel and 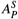 closely matched the GT, whereas FSL-MRS maps appeared noisier and exhibited regional systematic deviations (e.g., in HDO and Glc maps). Accordingly, very good correlation with the GT was achieved by 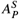 and LCModel (PCC ≥ 0.95). FSL-MRS fits showed good or very good correlation with the GT (PCC: 0.84 −0.97). The best correlations (PCC = 0.99) and SSIMs (≥ 0.99) for Glc, Glx, and HDO were achieved by 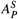 .

Relative difference maps confirmed that 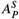 yielded the smallest deviations from the GT with the smallest RMSEs for Glc, Glx, and HDO, followed by LCModel, which showed slightly larger relative differences and more pronounced regional systematic deviation from the GT (e.g., in the HDO maps). For Lac, all methods exhibited larger relative differences. LCModel strongly overestimated Lac concentrations, while 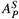 underestimated them. FSL-MRS showed regional systematic deviations in both direction. The best SSIM, PCC, and RMSE values for Lac were achieved by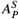.

#### 3.3.2. Model Constraint Parameters

DAE instance 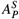 achieved good to excellent agreement with the GT model constraint parameters for Glc and HDO (SSIM: 0.90 −0.99, PCC: 0.76 −0.98), and moderate to good agreement for Glx (SSIM: 0.84 −0.92, PCC: 0.58 −0.92). Across all metabolites and metrics, 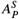 yielded strictly better values than the two reference methods. LCModel consistently outperformed FSL-MRS, except for Glx, where FSL-MRS achieved higher SSIMs.

## 4. Discussion

In this work, we presented a DAE for 2D fitting of *in vivo* dynamic DMI data, addressing the substantial time requirements associated with spectral quantification using classical MRS analysis tools. The proposed method enables quantification of whole-brain datasets within less than a second and supports fitting arbitrary parametric functions to describe the time evolution of metabolite concentrations. It incorporates prior physical knowledge through a linear combination model closely related to LCModel’s signal model. The autoencoder structure enables self-supervised training, avoiding the problem of unavailable GT for *in vivo* data. Although other autoencoders for fast spectral fitting of MRSI data have been proposed in recent years (Giuffrida et al., 2025; Gurbani et al., 2019; Shamaei et al., 2023, Shamaei et al., 2025), the proposed DAE is the only one developed for dynamic DMI data, and the only one that supports 2D fitting.

While most fitting tools assume pure Lorentzian or Voigt lineshapes (Borbath et al., 2021; Gurbani et al., 2019; Turco et al., 2024; Giuffrida et al., 2025; Shamaei et al., 2023, Shamaei et al., 2025; Clarke et al., 2021, Clarke et al., 2024), LCModel employs a nearly model-free, regularized lineshape kernel to account for various factors influencing the lineshape, such as *B*_0_ inhomogeneities (Provencher, 1993). We adopted this model-free lineshape kernel in our DAE to avoid systematic quantification biases caused by imperfect lineshapes.

Validation of the proposed DAE against 1D fitting based on LCModel showed excellent agreement on *in vivo* data for high-SNR metabolites (Glc, Glx, and HDO), preserving spatial structures in the metabolite maps, yielding low bias, and outperforming FSL-MRS across all evaluation metrics (SSIM, PCC, and Bland-Altman metrics) by large margin. For the low-SNR Lac signal, agreement was lower, with both FSL-MRS and the proposed DAE estimating approximately 50% lower signal intensities compared to LCModel on average. However, evaluation on synthetic data revealed that LCModel systematically overestimated Lac by 41% on average across both subjects from the synthetic test dataset, which may partly explain the discrepancies observed *in vivo*. This is consistent with findings by Borbath et al. (2021) on LCModel’s bias for 9.4T proton MRS data.

Quantitative evaluation using synthetic datasets showed strong agreement between the GT and metabolite concentrations fitted by the DAE. The fits of the model constraint parameters aligned well with expected metabolite concentration dynamics (e.g., shift between the signal increase of Glc and Glx), and strong agreement with the GT was found for most parameters. The proposed DAE outperformed both reference methods across all evaluation metrics for the fits of the model constraint parameters and metabolite concentrations. However, agreement on the model constraint parameters—especially for Glx—was lower than for metabolite concentrations, presumably due to poor identifiability of the logistic growth function when the spectral time courses lack full sigmoidal coverage along the temporal axis.

Despite strong similarities to LCModel in terms of signal model and cost function, the DAE outperformed LCModel on synthetic data. This may happen because of key modifications, such as using the full complex signal (rather than just the real part) for the reconstruction loss (Sokolenko et al., 2019), and exploiting the advantages of 2D fitting (Tal, 2023; Frese et al., 2024; Clarke et al., 2024). Furthermore, the large number of training samples seen by the DAE during training and tuning hyperparameters specifically for the application to dynamic DMI data likely contributed to superior performance. In contrast, FSL-MRS does not support free lineshape kernels but uses Lorentzian and Gaussian lineshapes, and models the baseline with complex polynomials instead of cubic B-splines, which might have contributed to the poorer performance on synthetic and *in vivo* data, despite implementing 2D fitting. Our evaluation showed that the proposed DAE reduced fitting times by more than three orders of magnitude to approximately 0.5 s for a whole-brain dataset compared to tens of minutes using conventional fitting tools.

### 4.1. Limitations and Outlook

Due to the lack of large dynamic DMI data sets, the number of subjects available for training, testing and validating the proposed DAE was limited. To better assess generalizability—particularly in the context of impaired glucose metabolism—additional data from a more diverse cohort of study participants would be valuable. However, we assume that a relatively small number of subjects suffices for training the proposed DAE, since a single dynamic DMI dataset contains roughly 2000 spectral time courses, and (Shamaei et al., 2023) showed that approximately 10^4^ training samples were adequate for a similar architecture trained on non-dynamic proton MRS data. Recent work by Shamaei et al. (2025) demonstrated that DAEs can provide uncertainty estimates with minimal computational overhead, which is a promising direction for extending our model. Currently, the model supports only model constraints expressible as closed-form parametric functions, limiting its applicability to more complex metabolic models that go beyond the heuristic model constraints used in this work (Lanz et al., 2013).

The main technical challenge in including metabolic models expressed as ordinary differential equations (ODEs) is backpropagating through (black box) ODE-solvers. However, this can be addressed using the adjoint sensitivity method (Pontryagin, 2018), as shown by Chen et al. (2018) in the context of Neural ODEs, defining a direction for future work on DL-based 2D fitting. In a broader context, this work suggests that DAE-based approaches are promising for fast spectral fitting in more than one dimension, which opens new perspectives to develop advanced DL-based fitting methods, e.g., by using convolutional layers whose kernels extend along the spatial dimensions, allowing the network to exploit spatial context, and by incorporating prior knowledge via spatial regularization based on additional loss terms or splines (Kelm et al., 2012).

## 5. Conclusion

Fast and accurate metabolite quantification is essential for the clinical translation of dynamic DMI. DL-based methods such as DAEs have shown great potential for significantly accelerating spectral fitting without compromising accuracy.

In this work, we proposed and benchmarked the first DAE developed specifically for spectral-temporal fitting of dynamic DMI data, enabling metabolite concentration and model constraint parameter fits for whole-brain datasets in under one second, leveraging benefits of 2D fitting for the fit quality. The approach enables self-supervised learning, overcoming the lack of GT for *in vivo* MRSI data. We demonstrated on *in vivo* and synthetic data that our model matches or surpasses LCModel and the FSL-MRS 2D fitting tool in terms of fitting accuracy. We believe that the proposed method offers a valuable step toward fast and robust post-processing pipelines for dynamic DMI, facilitating future clinical applications.

## Data Availability

The source code used to define and train the DAE model is publicly available at https://github.com/Osburg/dynamic_fitting. Data generated during this study (e.g., metabolite quantifications, basissets, scripts for figure generation), are available from the corresponding author upon reasonable request for non-commercial research purposes.

## Informed Consent

Written informed consent was obtained from all participants that underwent DMI scans for the acquisition of the in-vivo datasets used in this study.

## Declaration of generative AI in the writing process

During the preparation of this work the authors used GPT-4o in order to improve the readability and language of the manuscript. After using this tool/service, the authors reviewed and edited the content as needed and take full responsibility for the content of the published article.

## CRediT Authorship Contribution Statement

**Aaron P. Osburg:** Conceptualization, Data Curation, Formal analysis, Investigation, Methodology, Software, Validation, Visualization, Writing - Original Draft, Writing - Review & Editing. **Amirmohammed Shamaei:** Conceptualization, Methodology, Software, Writing - Review & Editing. **Anna Duguid:** Conceptualization, Writing - Review & Editing. **Bernhard Strasser:** Conceptualization, Data Curation, Writing - Review & Editing. **Fabian Niess:** Conceptualization, Investigation, Writing - Review & Editing. **Georg Langs:** Project administration, Writing - Review & Editing. **Hauke Fischer:** Conceptualization, Writing - Review & Editing. **Lukas Hingerl:** Conceptualization, Writing - Review & Editing. **Sabina Frese:** Conceptualization, Writing - Review & Editing. **William T. Clarke:** Software, Writing - Review & Editing. **Stanislav Motyka:** Conceptualization, Methodology, Project administration, Supervision, Writing - Review & Editing. **Viola Bader:** Conceptualization, Writing - Review & Editing. **Wolfgang Bogner:** Conceptualization, Methodology, Project administration, Resources, Supervision, Writing - Review & Editing.

## Acknowledgements

We gratefully acknowledge the financial support by the Christian Doppler Laboratory for MR Imaging Biomarkers (BIOMAK), the Austrian Federal Ministry for Digital and Economic Affairs, and the National Foundation for Research, Technology and Development and the Austrian Science Fund. This research was funded in whole or in part by the Austrian Science Fund (FWF) [10.55776/KLI1106/P34198]. For open access purposes, the author has applied a CC BY public copyright license to any author accepted manuscript version arising from this submission. Funded by the European Union (ERC, GLUCO-SCAN, 101088351). Views and opinions expressed are however those of the authors only and do not necessarily reflect those of the European Union or the European Research Council Executive Agency. Neither the European Union nor the granting authority can be held responsible for them. William T. Clarke is funded by Wellcome [225924/Z/22/Z].

---

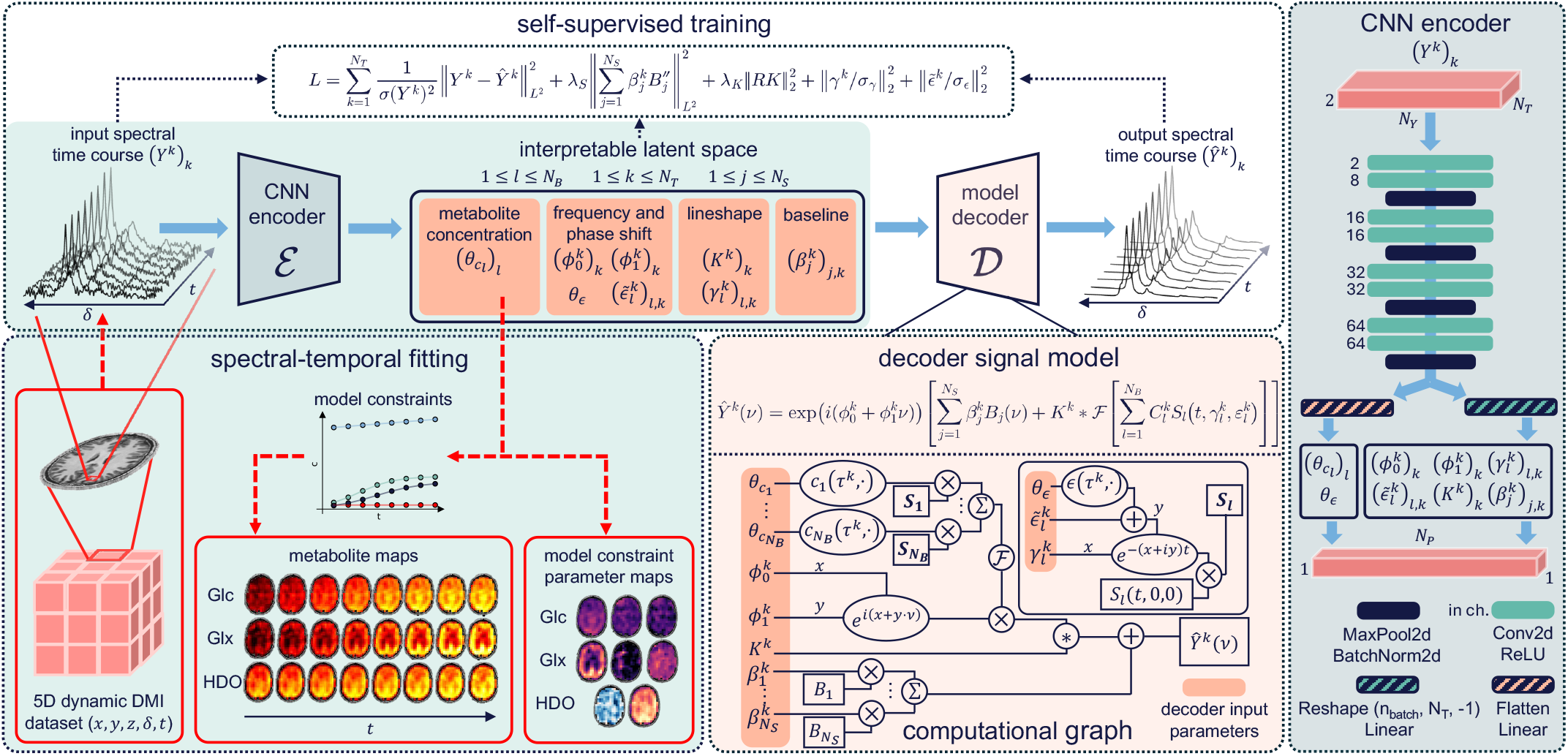

