## Supporting Information for "A Deep Autoencoder for Fast Spectral-Temporal Fitting of Dynamic Deuterium Metabolic Imaging Data at 7T"

**Supporting Information Figure S1:** (A): Metabolite maps of the three major metabolite signals (Glc, Glx, HDO) across all eight time points for the first subject from the *in vivo* test dataset, fitted by DAE instance $A_{I}^{I}$ and LCModel (without model constraint fits along the temporal axis). (B): Exemplary spectral time course with corresponding DAE fits and residuals. The index k indicates the position of each spectrum in the time series, and metabolite labels identify the spectral peaks. (C): T1-weighted anatomical image with a red box indicating the voxel location of the time course and a red line marking the position of the presented metabolite maps.

**Supporting Information Figure S2:** Top row: Signal intensities for Glc, Glx, HDO, and Lac fitted by $A_{I}^{I}$(markers), along with the corresponding model constraint fits (solid lines) for three representative voxels from the test subjects. The x-axis indicates the repetition index k. Bottom row: Histograms of the residuals $\varepsilon$computed across all six subjects and time points. Visual inspection suggests that the residuals for Glc, Glx, and HDO are approximately Gaussian distributed.

**Supporting Information Figure S3:** (A): Metabolite maps of the three major metabolite signals (Glc, Glx, HDO) across all eight time points for the second subject from the *in vivo* test dataset, fitted by DAE instance $A_{P}^{I}$ and the reference methods LCModel and FSL-MRS. (B): Example spectral time course with corresponding DAE fits and residuals. The index k indicates the position of each spectrum in the time series, and metabolite labels identify the spectral peaks. (C): T1-weighted anatomical image with a red box indicating the voxel location of the time course and a red line marking the position of the presented metabolite maps.

**Supporting Information Figure S4:** Lac maps for both subjects of the test dataset fitted by $A_{P}^{I}$.

**Supporting Information Figure S5:** (A): Metabolite maps of the three major metabolite signals (Glc, Glx, HDO) for the second subject of the *in vivo* test dataset and all eight time points fitted by DAE instance $A_{P}^{I}$. For each metabolite, a sagittal (first row), coronal (second row) and axial (third row) map is shown. (B): Corresponding maps of the fitted model constraint parameters.

**Supporting Information Figure S6:** Metabolite maps of the second subject of the synthetic test dataset for the time points $k\in\{2,5,8\}$ fitted by DAE instance $A_{P}^{S}$, LCModel, and FSL-MRS. The GT maps are shown in the right column. To the right of each fitted map, a map of the relative differences $\Delta c_{rel}=\left( c_{fit}-c_{GT} \right)/c_{GT}$ (where $c_{fit}$ is the fitted and $c_{GT}$ the GT concentration) to the GT is shown.

**Supporting Information Figure S7:** Correlation plot for the metabolite concentrations and model constraints for Glx fitted by DAE instance $A_{P}^{S}$, LCModel, and FSL-MRS versus GT for both subjects of the synthetic test dataset. Point distributions are visualized as density maps, with darker red indicating higher density. 25 randomly sampled data points per subject are overlaid as markers.

**Supporting Information Figure S8:** Correlation plot for the metabolite concentrations and model constraints for HDO fitted by DAE instance $A_{P}^{S}$, LCModel, and FSL-MRS versus GT for both subjects of the synthetic test dataset. Point distributions are visualized as density maps, with darker red indicating higher density. 25 randomly sampled data points per subject are overlaid as markers.

**Supporting Information Table S1:** Model and training pipeline configuration for each instance of the proposed DAE.

**Supporting Information Table S2:** Summary of the CNN encoder network for all instances of the proposed DAE. Conv2d: layer applying a two-dimensional convolution over an input signal composed of several input planes, ReLU: Rectified Linear Unit, Flatten: Collapses all dimensions but the batch dimension of an input tensor into a single dimension.

**Supporting Information Table S3:** Whole-brain processing times for the in-vivo ($A_{I}^{I}$, $A_{P}^{I}$, FSL-MRS, LCModel) and synthetic ($A_{P}^{S}$) test subjects for the DAE instances, FSL-MRS, and LCModel.

**Supporting Information Table S4:** SSIM, PCC, and Bland-Altman metrics for the metabolite concentration fits expressed as percentages of the mean of both measurements for DAE instance $A_{P}^{I}$ and FSL-MRS versus LCModel. The presented values are averages calculated over both subjects from the in-vivo test dataset.

**Supporting Information Table S5:** SSIM, PCC, and RMSE values for the metabolite concentration fits from $A_{P}^{S}$, LCModel and FSL-MRS compared to the GT for both subjects from the synthetic test dataset.

**Supporting Information Table S6:** SSIM, PCC, and RMSE values for the model constraint parameters fitted by $A_{P}^{S}$, LCModel and FSL-MRS compared to the GT for both subjects from the synthetic test dataset. Extreme outliers occurring in FSL-MRS and 1D-fitting were removed using the modified z-score method with a threshold value of 5 before calculating the metrics [1].

**Supporting Information Text S1:** Least-squares fitting methods were used to fit parametric model constraints to metabolite concentration estimates. All least-squares fits were carried out using the SciPy 1.13.1 implementation *scipy.optimize.curve_fit()* [2]. If this method failed for a given voxel, a second least squares fit using *scipy.optimize.minimize()* and the cost function

$$C\left( \theta\right)=\sum_{i=1}^{N_{T}} \left( y_{i}-f\left( t_{i},\theta\right) \right)^{2}$$

Is performed. Here, $y_{i}$ are the metabolite concentration estimates, $f$ is the model constraint, $t_{i}$ is the i-th time point and $\theta$ are the model constraint parameters. The following initial guesses were used for the model constraints applied in this work:

- Logistic function: $c_{max,0}=\frac{1}{N_{T}}\sum_{i=1}^{N_{T}} y_{i}, v_{0}=1, t_{0,0}=0$
- Linear function: $c_{0,0}=y_{1}, v_{0}=\frac{y_{N_{T}}-y_{1}}{t_{N_{T}}-t_{1}}$
- Constant function: $c_{0,0}\frac{1}{N_{T}}\sum_{i=1}^{N_{T}} y_{i}$

**Supporting Information Text S2:** The following list contains the fitting option settings and model definitions for dynamic fitting with FSL-MRS, an exemplary LCModel configuration file for one voxel and time point, and the fit parameters per spectral time course for the DAEs $A_{P}^{I}$ and $A_{P}^{S}$, FSL-MRS, and LCModel.

- LCModel configuration:

$LCMODL

OWNER='<owner>'

key = 210387309

Title='<name>’

HZPPPM=4.562554e+01, DELTAT=2.631600e-03, NUNFIL=96

FILBAS='<basisset_path>'

DOREFS(1) = F

DOREFS(2) = T

NREFPK(1) = 1

PPMREF(1,1) = 4.8

SHIFMN(1) = -0.3

SHIFMX(1) = 0.3

WSMET = 'DSS'

WSPPM = 0.0

N1HMET = 9

SUBBAS = T

NEACH = 99

WDLINE(6) = 0.0

PPMST = 8.0

PPMEND = 1

DEGZER = 0

DEGPPM = 0

SDDEGZ = 999

SDDEGP = 15

NSIMUL = 0

NOMIT = 2

CHOMIT(1) = 'inv_gl'

CHOMIT(2) = 'inv_wa'

NCALIB = 4

CHCALI(1) = 'water'

CHCALI(2) = 'Glx'

CHCALI(3) = 'Glc'

CHCALI(4) = 'Lac'

LTABLE = 7

LCSV = 0

LCOORD = 9

FILTAB='<output_path_table>'

FILCOO='<output_path_coord>'

FILRAW='<output_path_raw>'

FILPS='<output_path_ps>'

$END

- FSL-MRS model definition:

\# ------------------------------------------------------------------------

\# User file for defining a model

\# Parameter behaviour

\# 'variable' : one per time point

\# 'fixed'	: same for all time points

\# 'dynamic' : model-based change across time points

\# Each parameter of the spectral fitting gets a specific behaviour

\# The default behaviour is 'fixed'

Parameters = {

"conc": {

"other": "fixed",

"Glc": {

"dynamic": "model_sigmoid",

"params": ["amplitude_glc", "scaling_glc", "time_shift_glc"],

},

"Glx": {

"dynamic": "model_sigmoid",

"params": ["amplitude_glx", "scaling_glx", "time_shift_glx"],

},

"water": {"dynamic": "model_linear", "params": ["offset_water", "slope_water"]},

},

}

\# Optionally define bounds on the parameters

Bounds = {

"amplitude_glc": (0, None),

"scaling_glc": (0, None),

"time_shift_glc": (None, None),

"amplitude_glx": (0, None),

"scaling_glx": (0, None),

"time_shift_glx": (None, None),

"offset_water": (None, None),

"slope_water": (None, None),

"gamma": (0, None),

"sigma": (0, None),

}

\# Dynamic models here

\# These define how the parameters of the dynamic models change as a function

\# of the time variable (in dwMRS, that is the bvalue)

from numpy import asarray, exp, ones_like

def sigmoid(x):

return 1.0 / (1.0 + exp(-x))

def sigmoid_grad(x):

return sigmoid(x) * (1 - sigmoid(x))

def model_sigmoid(p, t):

\# p = [amp, scaling, time_shift]

return p[0] * sigmoid(10.0 * p[1] * (t - p[2]))

def model_linear(p, t):

\# p = [amp,slope]

return p[0] + p[1] * t

\# ------------------------------------------------------------------------

\# Gradients

def model_sigmoid_grad(p, t):

\# p = [amp, scaling, time_shift]

g0 = sigmoid(10.0 * p[1] * (t - p[2]))

g1 = 10.0 * p[0] * (t - p[2]) * sigmoid_grad(10.0 * p[1] * (t - p[2]))

g2 = -10.0 * p[0] * p[1] * sigmoid_grad(10.0 * p[1] * (t - p[2]))

return asarray([g0, g1, g2], dtype=object)

def model_linear_grad(p, t):

\# p = [amp,slope]

g0 = ones_like(t)

g1 = t

return asarray([g0, g1], dtype=object)

- FSL-MRS fitting option settings:

fsl_mrs_proc phase --file csi_dyn_data_2H.nii --ppm 4 5 --output processed_alltp_shift --filename phased

fsl_dynmrs --data processed_alltp_shift/phased.nii.gz --basis fid_2ms_glx_shift --output fsl_mrs --dyn_config resources/dmi_model.py --time_var resources/dmi_time_real_8tp.csv --ppmlim 0 6 --baseline_order 0 --overwrite --verbose --report --spatial-mask mask.nii --fslsub-queue veryshort.q --save-fit

- The number of fit parameters per spectral time course were 16 (FSL-MRS), 289 (DAE), and 257 (LCModel; 31 fit parameters per time point for the fit along the spectral axis; 9 fit parameters for the fit along the temporal axis).

**Supporting Information Text S3:** The performance metrics (PCC, RMSE, SSIM) were calculated as follows:

- RMSE: The RMSE between two maps $\left( X_{i} \right)_{i}$ and $\left( Y_{i} \right)_{i}$ with linear indices $1\leq i\leq N$ was computed as

$$RMSE=\sqrt{\frac{1}{N}\sum_{i=1}^{N} \left( X_{i}-Y_{i} \right)^{2}}$$

- PCC: Calculated with the SciPy 1.13.1 implementation *scipy.stats.pearsonr()* using default values for the optional arguments [2].
- SSIM: Calculated with the SciKit-Image 0.24.0 implementation *skimage.metrics.structural_similarity.ssim()* [3]. If GT was involved in the comparison, the data range of the GT map was used as *data_range* argument, otherwise the data range of the LCModel map was used. Default values were used for all other optional arguments.

Extreme outliers were filtered using modified z-score outlier detection with a threshold of 5 [1]. A voxel was detected as outlier if the modified z-score exceeded the threshold for either GT, the respective DAE instance, LCModel or FSL-MRS. For SSIM computations, outliers were replaced with zeros. For computing the PCC and RMSE, outliers were disregarded.

**Supporting Information Text S4:** The global scaling factors $\lambda$ for the concentrations and model constraint parameters fitted by LCModel and FSL-MRS were chosen to minimize the RMSE

$$RMSE=\frac{1}{N}\left\| c_{GT}-{\lambda c}_{LCModel/FSL-MRS} \right\|_{2}^{2}$$

between the GT and fitted HDO concentration levels. Here, $c_{GT}$ and $c_{LCModel/FSL-MRS}$represent vectors containing the HDO concentrations (GT, fitted by LCModel or FSL-MRS, respectively) from all synthetic datasets and time points, and $N$ is the number of vector components. The RMSE is minimal, if

$$\lambda=\frac{\langle c_{GT}, c_{LCModel/FSL-MRS}\rangle}{\left\langle c_{LCModel/FSL-MRS},c_{LCModel/FSL-MRS} \right\rangle},$$

where $\langle\cdot,\cdot\rangle$ denotes the dot product.

| **Configuration Parameter** | $\boldsymbol{A}_{\boldsymbol{I}}^{\boldsymbol{I}}$ | $\boldsymbol{A}_{\boldsymbol{P}}^{\boldsymbol{I}}\boldsymbol{/}\boldsymbol{A}_{\boldsymbol{P}}^{\boldsymbol{S}}$ |
| --- | --- | --- |
| Latent space variable scaling | basis signals: 10^3^  frequency shift: 1  Lorentzian damping: 10  Baseline spline coefficients: 1  0^th^ order phase shift: 1  1^st^ order phase shift: 1 | basis signals: 10^3^  frequency shift: 1  Lorentzian damping: 10  Baseline spline coefficients: 1  0^th^ order phase shift: 1  1^st^ order phase shift: 1 |
| Model constraint | Glc: None (non-dynamic fit)  Glx: None (non-dynamic fit)  HDO: None (non-dynamic fit)  Lac: None (non-dynamic fit)  frequency shift: None (non-dynamic fit) | Glc: logistic function  Glx: logistic function  HDO: linear  Lac: constant  frequency shift: None |
| Spectral interval | 0.7 ppm – 6.7 ppm | 0.7 ppm – 6.7 ppm |
| Lineshape kernel size | 0.49 ppm | 0.49 ppm |
| $\lambda_{S}$ | 10^10^ | 10^10^ |
| $\lambda_{K}$ | 30 | 30 |
| $\sigma_{\gamma}$ | 1 | 1 |
| $\sigma_{\epsilon}$ | 0.2 | 0.2 |
| Number of spline knots | 4 | 4 |
| Batch size | 256 | 256 |
| Learning rate | 0.002 | 0.002 |
| Weight decay | 0.001 | 0.001 |
| Learning rate reduction factor | 0.5 | 0.5 |
| Epochs for learning rate reduction | 20, 50, 100, 200, 300, 400, 600 | 20, 50, 100, 200, 300, 400, 600 |
| Float32 matrix multiplication precision | “highest” | “highest” |
| Floating point format | Float32/complex64 | Float32/complex64 |

**Supporting Information Table S1:** Model and training pipeline configuration for each instance of the proposed DAE.

| **Model Instance** | **Layer** | **Input Shape** | **Output Shape** | **Number of Parameters** |
| --- | --- | --- | --- | --- |
| $A_{I}^{I}$ | Conv2d-1 | (B, 2, 8, 184) | (B, 8, 8, 184) | 248 |
|  | ReLU-1 | (B, 8, 8, 184) | (B, 8, 8, 184) | 0 |
|  | Conv2d-2 | (B, 8, 8, 184) | (B, 16, 8, 184) | 1.936 |
|  | ReLU-2 | (B, 16, 8, 184) | (B, 16, 8, 184) | 0 |
|  | MaxPool2d-1 | (B, 16, 8, 184) | (B, 16, 4, 92) | 0 |
|  | BatchNorm2d-1 | (B, 16, 4, 92) | (B, 16, 4, 92) | 32 |
|  | Conv2d-3 | (B, 16, 4, 92) | (B, 16, 4, 92) | 3.856 |
|  | ReLU-3 | (B, 16, 4,92) | (B, 16, 4, 92) | 0 |
|  | Conv2d-4 | (B, 16, 4,92) | (B, 32, 4, 92) | 7.712 |
|  | ReLU-4 | (B, 32, 4, 92) | (B, 32, 4, 92) | 0 |
|  | MaxPool2d-2 | (B, 32, 4, 92) | (B, 32, 2, 46) | 0 |
|  | BatchNorm2d-2 | (B, 32, 2, 46) | (B, 32, 2, 46) | 64 |
|  | Conv2d-5 | (B, 32, 2, 46) | (B, 32, 2, 46) | 15.392 |
|  | ReLU-5 | (B, 32, 2, 46) | (B, 32, 2, 46) | 0 |
|  | Conv2d-6 | (B, 32, 2, 46) | (B, 64, 2,46) | 30784 |
|  | ReLU-6 | (B, 64, 2, 46) | (B, 64, 2,46) | 0 |
|  | MaxPool2d-3 | (B, 64, 2, 23) | (B, 64, 2, 23) | 0 |
|  | BatchNorm2d-3 | (B, 64, 2, 23) | (B, 64, 2, 23) | 128 |
|  | Conv2d-7 | (B, 64, 2, 23) | (B, 64, 2, 12) | 61504 |
|  | ReLU-7 | (B, 64, 2, 12) | (B, 64, 2, 12) | 0 |
|  | Conv2d-8 | (B, 64, 2, 12) | (B, 64, 2, 6) | 61504 |
|  | ReLU-8 | (B, 64, 2, 6) | (B, 64, 2, 6) | 0 |
|  | Linear-1 | (B, 8, 96) | (B, 8, 34) | 3298 |
|  | Linear-2 | (B, 768) | (B, 40) | 30760 |
| Total Number of Parameters | 217.218 |  |  |  |
| Total Estimated Model size | 0.869 MB |  |  |  |
| $A_{P}^{I}/A_{P}^{S}$ | Conv2d-1 | (B, 2, 8, 184) | (B, 8, 8, 184) | 248 |
|  | ReLU-1 | (B, 8, 8, 184) | (B, 8, 8, 184) | 0 |
|  | Conv2d-2 | (B, 8, 8, 184) | (B, 16, 8, 184) | 1.936 |
|  | ReLU-2 | (B, 16, 8, 184) | (B, 16, 8, 184) | 0 |
|  | MaxPool2d-1 | (B, 16, 8, 184) | (B, 16, 4, 92) | 0 |
|  | BatchNorm2d-1 | (B, 16, 4, 92) | (B, 16, 4, 92) | 32 |
|  | Conv2d-3 | (B, 16, 4, 92) | (B, 16, 4, 92) | 3.856 |
|  | ReLU-3 | (B, 16, 4,92) | (B, 16, 4, 92) | 0 |
|  | Conv2d-4 | (B, 16, 4,92) | (B, 32, 4, 92) | 7.712 |
|  | ReLU-4 | (B, 32, 4, 92) | (B, 32, 4, 92) | 0 |
|  | MaxPool2d-2 | (B, 32, 4, 92) | (B, 32, 2, 46) | 0 |
|  | BatchNorm2d-2 | (B, 32, 2, 46) | (B, 32, 2, 46) | 64 |
|  | Conv2d-5 | (B, 32, 2, 46) | (B, 32, 2, 46) | 15.392 |
|  | ReLU-5 | (B, 32, 2, 46) | (B, 32, 2, 46) | 0 |
|  | Conv2d-6 | (B, 32, 2, 46) | (B, 64, 2,46) | 30784 |
|  | ReLU-6 | (B, 64, 2, 46) | (B, 64, 2,46) | 0 |
|  | MaxPool2d-3 | (B, 64, 2, 23) | (B, 64, 2, 23) | 0 |
|  | BatchNorm2d-3 | (B, 64, 2, 23) | (B, 64, 2, 23) | 128 |
|  | Conv2d-7 | (B, 64, 2, 23) | (B, 64, 2, 12) | 61504 |
|  | ReLU-7 | (B, 64, 2, 12) | (B, 64, 2, 12) | 0 |
|  | Conv2d-8 | (B, 64, 2, 12) | (B, 64, 2, 6) | 61504 |
|  | ReLU-8 | (B, 64, 2, 6) | (B, 64, 2, 6) | 0 |
|  | Linear-1 | (B, 8, 96) | (B, 8, 34) | 3298 |
|  | Linear-2 | (B, 768) | (B,17) | 13073 |
| Total Number of Parameters | 199.531 |  |  |  |
| Total Estimated Model size | 0.798 MB |  |  |  |

**Supporting Information Table S2:** Summary of the CNN encoder network for all instances of the proposed DAE. Conv2d: layer applying a two-dimensional convolution over an input signal composed of several input planes, ReLU: Rectified Linear Unit, Flatten: Collapses all dimensions but the batch dimension of an input tensor into a single dimension. B: Size of the input tensor along the batch dimension. The models have no non-trainable parameters.

| **Method** | **Test subject 1 (1771 spectral time courses)** | **Test subject 2 (1902 spectral time courses)** | **Mean processing time per spectral time course** | **Mean processing time per subject** |
| --- | --- | --- | --- | --- |
| DAE $A_{P}^{I}$ | 0.51 s | 0.58 s | 0.29 ms | 0.55 s |
| DAE $A_{P}^{S}$ | 0.52 s | 0.52 s | 0.28 ms | 0.52 s |
| DAE $A_{I}^{I}$ | 0.51 s | 0.53 s | 0.28 ms | 0.52 s |
| FSL-MRS | 1184 s | 1220 s | 0.65 s | 1202 s |
| LCModel | 944 s | 1088 s | 0.55 s | 1016 s |

**Supporting Information Table S3:** Whole-brain processing times for the in-vivo ($A_{I}^{I}$, $A_{P}^{I}$, FSL-MRS, LCModel) and synthetic ($A_{P}^{S}$) test subjects for the DAE instances, FSL-MRS, and LCModel.

|  | | **Test Subject 1** | | | | **Test Subject 2** | | | |
| --- | --- | --- | --- | --- | --- | --- | --- | --- | --- |
| **Metabolite** | **Method** | **SSIM** | **PCC** | **Bias [%]** | **LoA [%]** | **SSIM** | **PCC** | **Bias [%]** | **LoA [%]** |
| **Glc** | $\boldsymbol{A}_{\boldsymbol{P}}^{\boldsymbol{I}}$ | **0.98** | **0.96** | **10** | **110** | **0.99** | **0.98** | **7.9** | **78** |
|  | **FSL-MRS** | 0.76 | 0.71 | 14 | 144 | 0.94 | 0.90 | 8.5 | 109 |
| **Glx** | $\boldsymbol{A}_{\boldsymbol{P}}^{\boldsymbol{I}}$ | **0.98** | **0.97** | **8.6** | **143** | **0.98** | **0.97** | **11** | **137** |
|  | **FSL-MRS** | 0.67 | 0.58 | 28 | 167 | 0.90 | 0.88 | 26 | 157 |
| **HDO** | $\boldsymbol{A}_{\boldsymbol{P}}^{\boldsymbol{I}}$ | **0.98** | **0.93** | **1.0** | **41** | **0.99** | **0.94** | **-0.10** | **26** |
|  | **FSL-MRS** | 0.73 | 0.27 | 2.0 | 86 | 0.91 | 0.60 | -4.5 | 54 |
| **Lac** | $\boldsymbol{A}_{\boldsymbol{P}}^{\boldsymbol{I}}$ | **0.70** | **0.69** | -41 | **219** | **0.68** | **0.71** | **-52** | **220** |
|  | **FSL-MRS** | 0.52 | 0.45 | **-38** | 278 | 0.67 | 0.71 | -58 | 262 |

**Supporting Information Table S4:** SSIM, PCC, and Bland-Altman metrics for the metabolite concentration fits expressed as percentages of the mean of both measurements for DAE instance $A_{P}^{I}$ and FSL-MRS versus LCModel. The presented values are averages calculated over both subjects from the in-vivo test dataset.

|  | | **Test Subject 1** | | | **Test Subject 2** | | |
| --- | --- | --- | --- | --- | --- | --- | --- |
| **Metabolite** | **Method** | **SSIM** | **PCC** | **RMSE [10^-2^ a.u.]** | **SSIM** | **PCC** | **RMSE [10^-2^ a.u.]** |
| **Glc** | $\boldsymbol{A}_{\boldsymbol{P}}^{\boldsymbol{S}}$ | **1.00** | **0.99** | **1.8** | **0.99** | **0.99** | **1.8** |
|  | **LCModel** | 0.96 | 0.97 | 3.7 | **0.99** | **0.99** | 2.8 |
|  | **FSL-MRS** | 0.95 | 0.92 | 5.0 | 0.97 | 0.96 | 4.1 |
| **Glx** | $\boldsymbol{A}_{\boldsymbol{P}}^{\boldsymbol{S}}$ | **0.99** | **0.99** | **2.8** | **0.99** | **0.99** | **2.7** |
|  | **LCModel** | 0.97 | 0.98 | 4.2 | 0.98 | 0.98 | 3.7 |
|  | **FSL-MRS** | 0.97 | 0.97 | 4.4 | 0.98 | 0.97 | 3.7 |
| **HDO** | $\boldsymbol{A}_{\boldsymbol{P}}^{\boldsymbol{S}}$ | **1.00** | **0.99** | **4.5** | **1.00** | **0.99** | **4.1** |
|  | **LCModel** | 0.96 | 0.95 | 12 | 0.99 | 0.96 | 7.5 |
|  | **FSL-MRS** | 0.95 | 0.84 | 18 | 0.96 | 0.85 | 14 |
| **Lac** | $\boldsymbol{A}_{\boldsymbol{P}}^{\boldsymbol{S}}$ | **0.86** | **0.87** | **2.0** | **0.88** | **0.89** | **1.8** |
|  | **LCModel** | 0.77 | 0.83 | 2.5 | 0.85 | 0.86 | 2.1 |
|  | **FSL-MRS** | 0.80 | 0.71 | 2.9 | 0.82 | 0.76 | 2.4 |

**Supporting Information Table S5:** SSIM, PCC, and RMSE values for the metabolite concentration fits from $A_{P}^{S}$, LCModel and FSL-MRS compared to the GT for both subjects from the synthetic test dataset.

|  | | | **Test Subject 1** | | | **Test Subject 2** | | |
| --- | --- | --- | --- | --- | --- | --- | --- | --- |
| **Metabolite** | **Parameter** | **Method** | **SSIM** | **PCC** | **RMSE [a.u.]** | **SSIM** | **PCC** | **RMSE**  **[a.u.]** |
| **Glc** | **c_max_ [a.u.]** | $\boldsymbol{A}_{\boldsymbol{P}}^{\boldsymbol{S}}$ | **0.99** | **0.98** | **0.019** | **0.99** | **0.98** | **0.021** |
|  |  | **LCModel** | 0.91 | 0.90 | 0.042 | 0.96 | 0.92 | 0.037 |
|  |  | **FSL-MRS** | 0.75 | 0.38 | 0.012 | 0.86 | 0.59 | 0.083 |
|  | **v [10^-3^ min^-1^]** | $\boldsymbol{A}_{\boldsymbol{P}}^{\boldsymbol{S}}$ | **0.95** | **0.74** | **1.1** | **0.95** | **0.76** | **0.99** |
|  |  | **LCModel** | 0.79 | 0.35 | 2.6 | 0.84 | 0.54 | 2.2 |
|  |  | **FSL-MRS** | 0.63 | 0.45 | 3.9 | 0.75 | 0.41 | 3.0 |
|  | **t_0_ [min]** | $\boldsymbol{A}_{\boldsymbol{P}}^{\boldsymbol{S}}$ | **0.98** | **0.89** | **1.7** | **0.98** | **0.87** | **1.6** |
|  |  | **LCModel** | 0.89 | 0.081 | 3.2 | 0.95 | 0.75 | 2.5 |
|  |  | **FSL-MRS** | 0.65 | 0.74 | 8.4 | 0.83 | 0.27 | 5.3 |
| **Glx** | **c_max_ [a.u.]** | $\boldsymbol{A}_{\boldsymbol{P}}^{\boldsymbol{S}}$ | **0.91** | **0.91** | **0.072** | **0.92** | **0.92** | **0.065** |
|  |  | **LCModel** | 0.76 | 0.80 | 0.11 | 0.78 | 0.82 | 0.098 |
|  |  | **FSL-MRS** | 0.85 | 0.78 | 0.12 | 0.86 | 0.83 | 0.091 |
|  | **v [10^-3^ min^-1^]** | $\boldsymbol{A}_{\boldsymbol{P}}^{\boldsymbol{S}}$ | **0.86** | **0.73** | **1.8** | **0.88** | **0.71** | **2.1** |
|  |  | **LCModel** | 0.69 | 0.55 | 3.7 | 0.71 | 0.58 | 4.1 |
|  |  | **FSL-MRS** | 0.74 | 0.59 | 3.1 | 0.77 | 0.63 | 3.3 |
|  | **t_0_ [min]** | $\boldsymbol{A}_{\boldsymbol{P}}^{\boldsymbol{S}}$ | **0.83** | **0.53** | **4.6** | **0.86** | **0.54** | **4.2** |
|  |  | **LCModel** | 0.74 | 0.45 | 7.3 | 0.81 | 0.44 | 7.0 |
|  |  | **FSL-MRS** | 0.76 | 0.31 | 7.4 | 0.83 | 0.46 | 5.7 |
| **HDO** | **c_0_ [a.u.]** | $\boldsymbol{A}_{\boldsymbol{P}}^{\boldsymbol{S}}$ | **0.91** | **0.89** | **0.14** | **0.90** | **0.88** | **0.14** |
|  |  | **LCModel** | 0.80 | 0.76 | 0.22 | 0.86 | 0.82 | 0.18 |
|  |  | **FSL-MRS** | 0.76 | 0.72 | 0.24 | 0.78 | 0.75 | 0.23 |
|  | **v [10^-2^ a.u./min]** | $\boldsymbol{A}_{\boldsymbol{P}}^{\boldsymbol{S}}$ | **0.99** | **0.97** | **0.069** | **0.99** | **0.97** | **0.067** |
|  |  | **LCModel** | 0.97 | 0.95 | 0.18 | 0.98 | 0.94 | 0.086 |
|  |  | **FSL-MRS** | 0.95 | 0.86 | 0.11 | 0.96 | 0.84 | 0.14 |

**Supporting Information Table S6:** SSIM, PCC, and RMSE values for the model constraint parameters fitted by $A_{P}^{S}$, LCModel and FSL-MRS compared to the GT for both subjects from the synthetic test dataset. Extreme outliers occurring in FSL-MRS and 1D-fitting were removed using the modified z-score method with a threshold value of 5 before calculating the metrics [1].

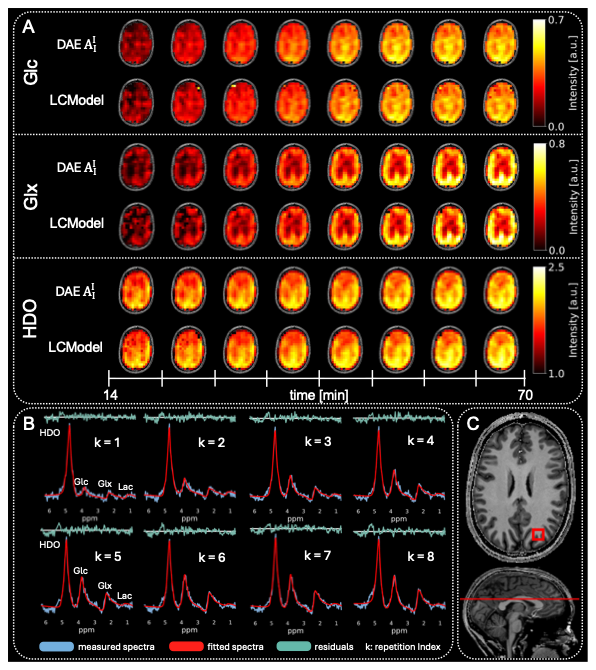

**Supporting Information Figure S1:** (A): Metabolite maps of the three major metabolite signals (Glc, Glx, HDO) across all eight time points for the first subject from the *in vivo* test dataset, fitted by DAE instance $A_{I}^{I}$ and LCModel (without model constraint fits along the temporal axis). (B): Exemplary spectral time course with corresponding DAE fits and residuals. The index k indicates the position of each spectrum in the time series, and metabolite labels identify the spectral peaks. (C): T1-weighted anatomical image with a red box indicating the voxel location of the time course and a red line marking the position of the presented metabolite maps.

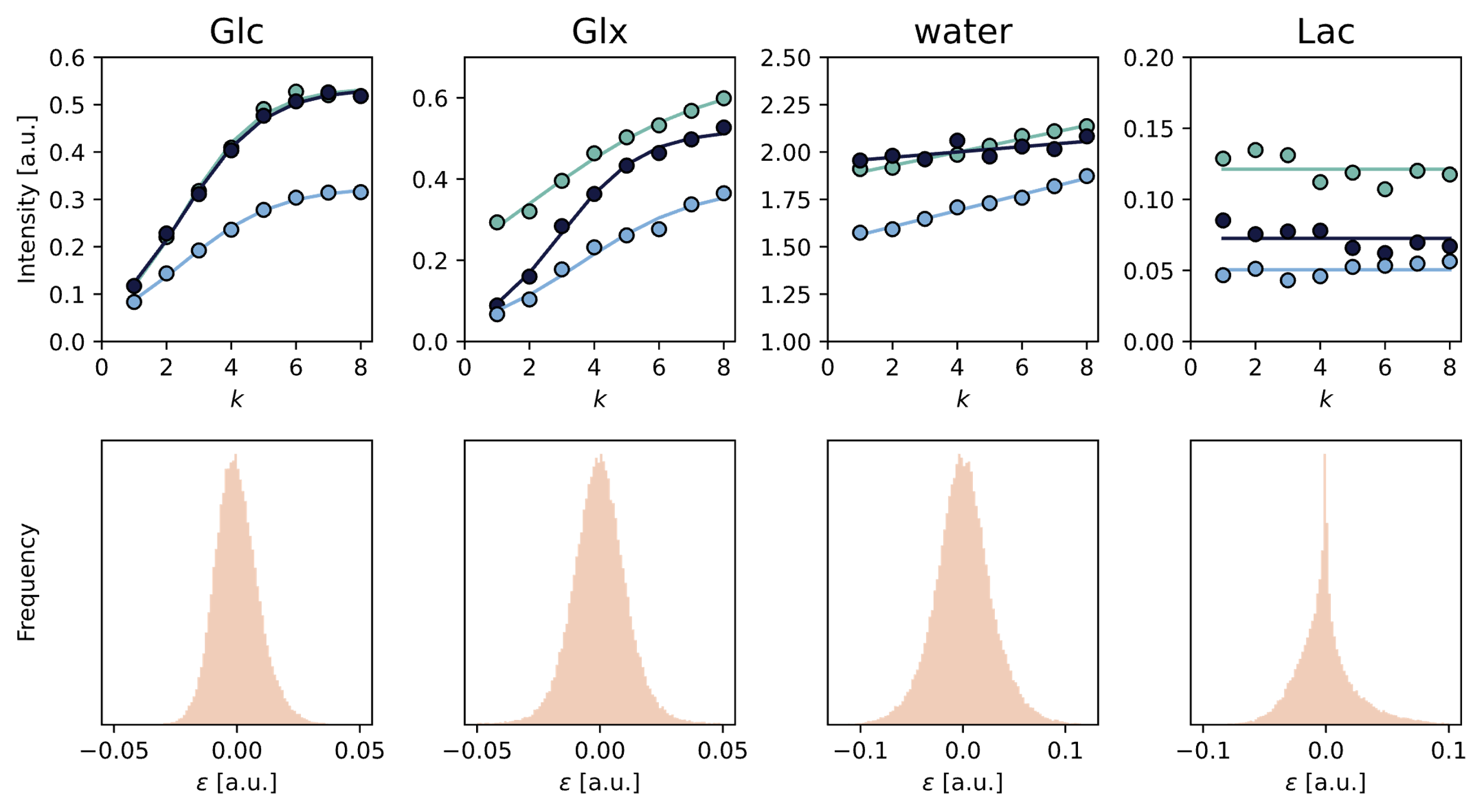

**Supporting Information Figure S2:** Top row: Signal intensities for Glc, Glx, HDO, and Lac fitted by $A_{I}^{I}$(markers), along with the corresponding model constraint fits (solid lines) for three representative voxels from the test subjects. The x-axis indicates the repetition index k. Bottom row: Histograms of the residuals $\varepsilon$computed across all six subjects and time points. Visual inspection suggests that the residuals for Glc, Glx, and HDO are approximately Gaussian distributed.

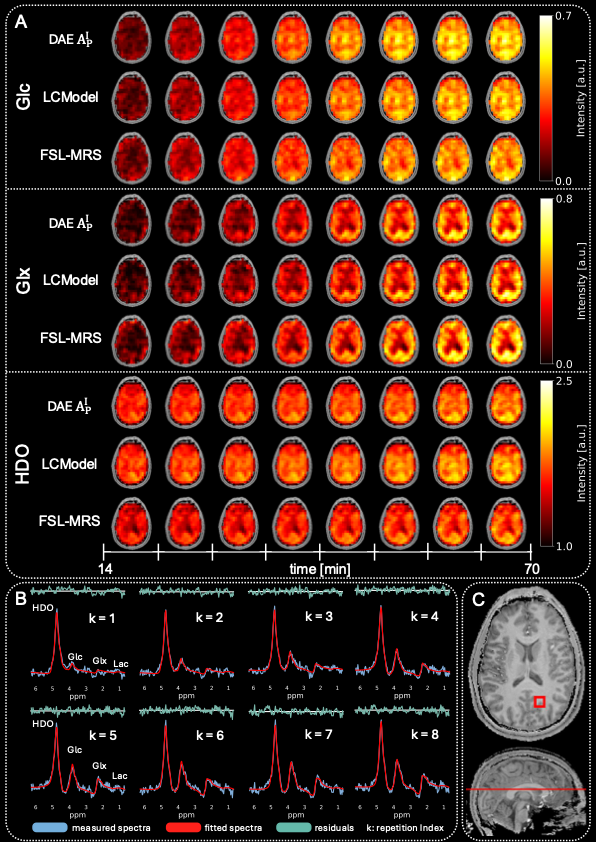

**Supporting Information Figure S3:** (A): Metabolite maps of the three major metabolite signals (Glc, Glx, HDO) across all eight time points for the second subject from the *in vivo* test dataset, fitted by DAE instance $A_{P}^{I}$ and the reference methods LCModel and FSL-MRS. (B): Example spectral time course with corresponding DAE fits and residuals. The index k indicates the position of each spectrum in the time series, and metabolite labels identify the spectral peaks. (C): T1-weighted anatomical image with a red box indicating the voxel location of the time course and a red line marking the position of the presented metabolite maps.

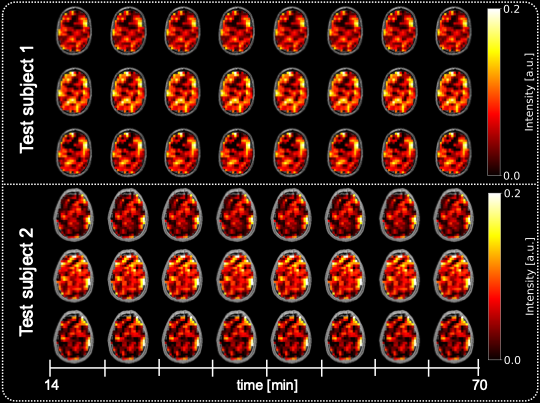

**Supporting Information Figure S4:** Lac maps for both subjects of the test dataset fitted by $A_{P}^{I}$.

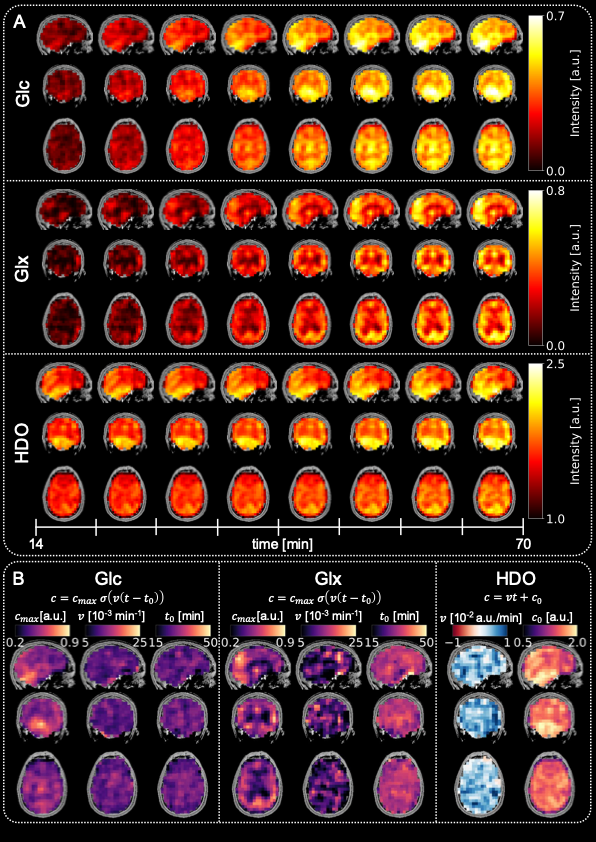

**Supporting Information Figure S5:** (A): Metabolite maps of the three major metabolite signals (Glc, Glx, HDO) for the second subject of the *in vivo* test dataset and all eight time points fitted by DAE instance $A_{P}^{I}$. For each metabolite, a sagittal (first row), coronal (second row) and axial (third row) map is shown. (B): Corresponding maps of the fitted model constraint parameters.

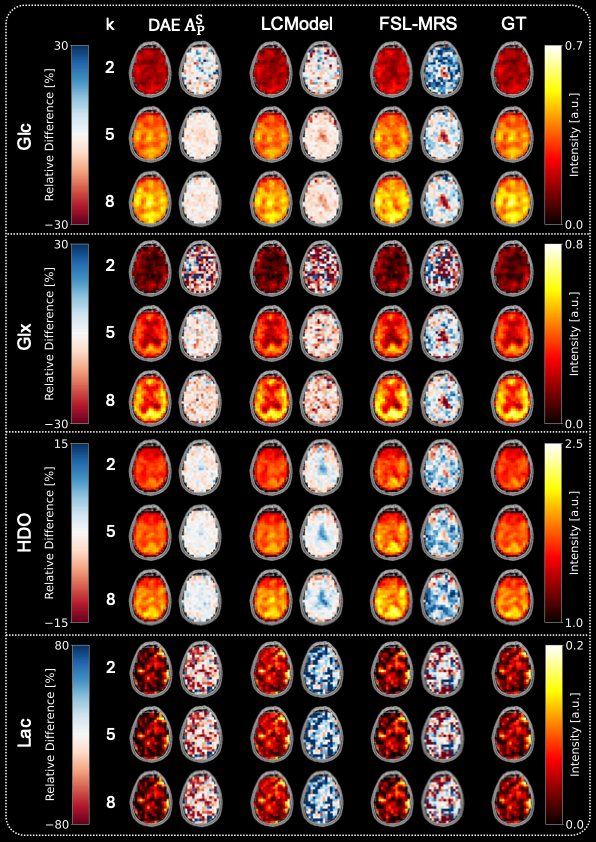

**Supporting Information Figure S6:** Metabolite maps of the second subject of the synthetic test dataset for the time points $k\in\{2,5,8\}$ fitted by DAE instance $A_{P}^{S}$, LCModel, and FSL-MRS. The GT maps are shown in the right column. To the right of each fitted map, a map of the relative differences $\Delta c_{rel}=\left( c_{fit}-c_{GT} \right)/c_{GT}$ (where $c_{fit}$ is the fitted and $c_{GT}$ the GT concentration) to the GT is shown.

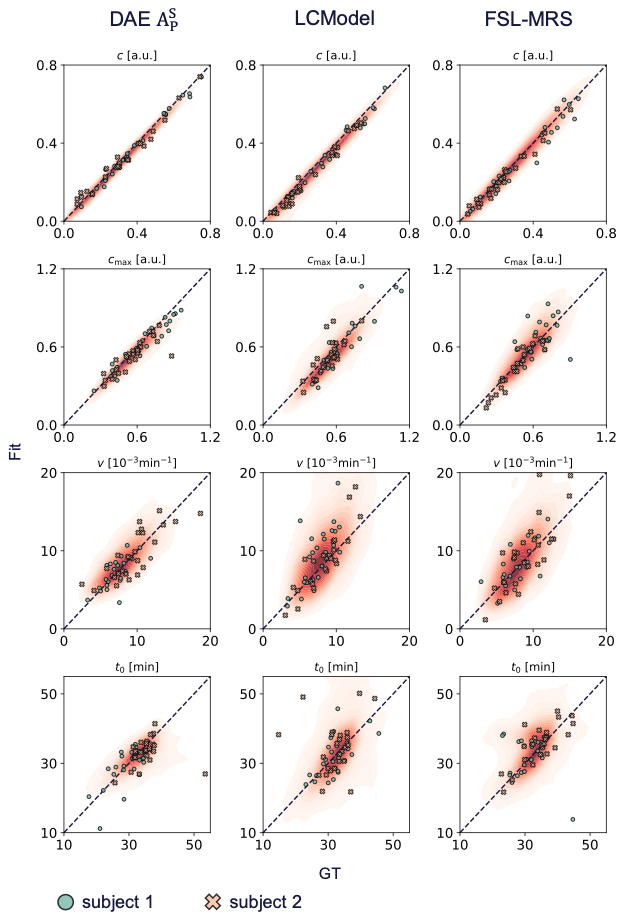

**Supporting Information Figure S7:** Correlation plot for the metabolite concentrations and model constraints for Glx fitted by DAE instance $A_{P}^{S}$, LCModel, and FSL-MRS versus GT for both subjects of the synthetic test dataset. Point distributions are visualized as density maps, with darker red indicating higher density. 25 randomly sampled data points per subject are overlaid as markers.

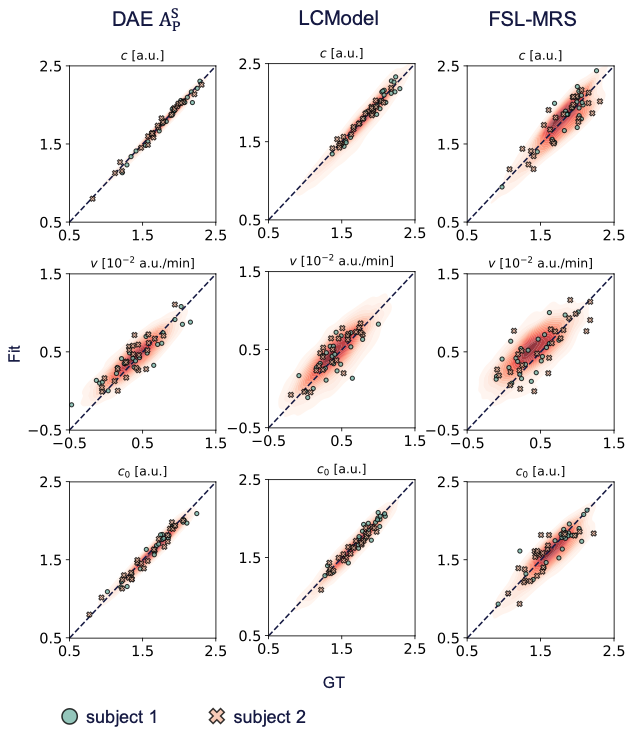

**Supporting Information Figure S8:** Correlation plot for the metabolite concentrations and model constraints for HDO fitted by DAE instance $A_{P}^{S}$, LCModel, and FSL-MRS versus GT for both subjects of the synthetic test dataset. Point distributions are visualized as density maps, with darker red indicating higher density. 25 randomly sampled data points per subject are overlaid as markers.

[3] van der Walt, S., Schönberger, J.L., Nunez-Iglesias, J., Boulogne, F., Warner, J.D., Yager, N., Gouillart, E., Yu, T.,

scikit-image contributors, 2014. scikit-image: image processing in python. PeerJ 2, e453.
